# NeuroFM: Toward Precision Neuroimaging with Foundation Models for Individualized Brain Health Estimation

**DOI:** 10.64898/2026.03.27.26349489

**Authors:** Austin Dibble, Connor Dalby, Michele Sevegnani, Alessio Fracasso, Donald M. Lyall, Monika Harvey, Michele Svanera, the Alzheimer’s Disease Neuroimaging Initiative, the Frontotemporal Lobar Degeneration Neuroimaging Initiative

## Abstract

Precision neuroimaging aims to deliver individualized assessments of brain health, yet a single structural MRI does not provide a scalable, multidimensional, quantitative summary of an individual’s current or future health. Existing approaches optimize task-specific objectives, yielding representations entangled with cohort- or disease-specific signals rather than capturing biologically grounded anatomical patterns. Here, we introduce NeuroFM, a foundation model trained exclusively on 100,000 healthy synthetic volumes to predict morphometric and demographic targets. Without exposure to disease-labelled data, NeuroFM organizes brain structure into population-level patterns encoding brain health differences. These representations transfer across neuroscience domains without adaptation and support simple linear readouts for clinical, cognitive, developmental, socio-behavioural, and image quality. Evaluated on 136,361 multi-cohort volumes, NeuroFM generalizes across domains and enables individual-level brain health profiling, estimating future dementia risk years before diagnosis. Together, these findings establish a disease-naïve foundation model for precision neuroimaging with potential to support quantitative brain health assessments across settings.

## 1 Main

Precision neuroimaging aims to quantify brain health at the individual level, enabling personalized assessments of neurological and psychiatric risk, resilience, and cognitive and behavioural capacity. Central to this aim is brain-health estimation: the inference of biologically meaningful indicators of structural integrity, functional capacity, and pathological alterations, as well as their age-expected trajectories and early deviations across lifespan. Brain-health estimation provides a conceptual framework for quantifying these properties across populations, whereas precision neuroimaging refers to the reliable realization of these brain health indicators at the individual level. Although brain health is inherently multidimensional, structural brain MRI provides an accessible and biologically grounded window into these processes, and strongly associates with cognitive and psychological outcomes [1–5]. However, despite its ubiquity in neuroscience and clinical research, structural MRI does not yet provide a practical framework for deriving individualized, quantitative estimates of brain health from a single volume, partly due to several long-standing methodological barriers [6, 7].

Structural MRI signals exhibit substantial biological and technical variability due to inter-individual differences in development and ageing, disease heterogeneity, and to variation in acquisition protocols and scanner hardware, making it challenging to learn representations that generalize reliably across tasks, populations, and imaging environments [8, 9]. Over the past two decades, several methodological paradigms have sought to overcome such challenges and to extract meaningful representations (Extended Fig. 1). Early approaches relied on dedicated morphometric features (i.e., hippocampal volume or cortical thickness) derived from pipelines such as FreeSurfer [10] or FSL [11], followed by classical machine-learning models, enabling interpretability but restricting analysis to a predefined set of anatomical measurements. Subsequent deep learning approaches learned features directly from MRI volumes [12, 13], improving predictive performance. However, these models typically optimize representations for specific downstream objectives, coupling the learned representation to task-specific labels, limiting generalizability and reuse across cohorts and domains [14–17]. Brain age gap modelling introduced an important conceptual shift toward biomarker-oriented neuroimaging by predicting deviations from age-expected trajectories; however, its scalar formulation and task-specific training collapse the multidimensional nature of brain health into a single measure [18–21]. Most recently, large-scale representation learning and neuroimaging foundation models have been introduced to mitigate these challenges through extensive pretraining, improving robustness and generalization [22, 23]. These approaches typically learn representations using generic self-supervised or proxy imaging objectives applied to real-world MRI datasets. These strategies produce transferable image features but the learned representations are not explicitly structured around biologically meaningful patterns of anatomical variation. Moreover, learning such representations directly from real-world MRI datasets is also challenging because anatomical variation is often entangled with disease effects, cohort composition, and acquisition variability [24, 25], as well as the limited availability of large, well-controlled normative datasets [1, 26]. As a result, the relationship between these representations and interpretable patterns of brain health remains indirect, limiting individualized inference across domains. This motivates a foundation-model paradigm explicitly designed for individualized brain-health estimation from structural MRI, bringing precision neuroimaging closer to practical application.

Here, we introduce NeuroFM, a brain MRI foundation model explicitly designed for structural precision neuroimaging. Rather than optimizing performance on downstream tasks, NeuroFM is pre-trained to learn biologically grounded representations of brain health by predicting a set of interpretable morphometric and demographic targets from structural MRI: age, sex, total brain volume (TBV), and ventricular volume. These targets were selected not as endpoints of interest, but as biologically meaningful proxies that compel the model to interrogate global and regional brain morphology, normative structural trajectories, and deviations associated with ageing and disease [1, 2, 27]. NeuroFM is trained exclusively on a large corpus of AI-generated human brain MRI volumes from LDM100k dataset (n=100,000) [28, 29]. These volumes represent healthy anatomical variation and avoid exposure to clinical labels or disease-specific datasets, enabling the learning of population-scale anatomical priors without reliance on real-world data. Crucially, the resulting model is deployed as a frozen feature extractor, with parameters that are fixed after pretraining, supporting individualized brain-health estimation across diverse domains using simple linear or logistic readouts, without requiring task-specific fine-tuning. As such, NeuroFM operationalizes precision neuroimaging as a property of the representation itself, enabling direct individualized inference from a single structural MRI volume.

We evaluated NeuroFM across a broad set of task datasets (136,361 volumes) on tasks spanning five neuroscience domains: clinical, cognitive, socio-behavioural, developmental, and image quality control. These domains were selected to systematically test the model’s generalizability, neuroscientific utility, and interpretability across diverse application settings. Lightweight models trained on NeuroFM features consistently outperformed task-specific approaches and existing representations reported in the literature, including both established baseline architectures and recent neuroimaging foundation models trained using generic computer-vision-style pretraining objectives. NeuroFM accurately identified neurodegenerative conditions, distinguished neurodevelopmental subtypes, and estimated MRI quality metrics, such as signal-to-noise ratio and motion. The model also generalized across MRI acquisition protocols with minimal adaptation and produced anatomically coherent attribution maps via layer-wise relevance propagation [4, 30, 31]. Finally, in a longitudinal case study using the ADNI dataset, NeuroFM achieved an individual-level prediction of future Alzheimer’s disease risk years before clinical diagnosis, demonstrating its potential for individualized brain-health forecasting.

NeuroFM provides a scalable, anatomically informed foundation model for precision neuroimaging, trained on disease-naïve synthetic MRI data to learn population-level representations of brain health, combining performance, sample efficiency, and interpretability. It supports integrated individual-level reporting across multiple patterns of brain health, including brain age gap, neurodegenerative risk estimates, cognitive and behavioural profiles, and MRI quality metrics. Together, these results establish a foundation-model paradigm for multidimensional, individualized brain-health estimation from structural MRI, enabling scalable precision neuroimaging applications. In practice, this framework could support standardized brain-health reporting, early risk stratification, and longitudinal monitoring from structural MRI in both research and clinical settings. Code, model weights, and documentation available at https://rocknroll87q.github.io/NeuroFM/.

## 2 Results

We developed NeuroFM, a foundation model for precision neuroimaging trained via supervised pretraining on large-scale synthetic brain MRI data. We then evaluated whether its learned representations encode biologically meaningful structure that generalizes across clinical, cognitive, and population settings. Rather than optimizing NeuroFM for task-specific performance, we designed downstream analyses as targeted tests of representational validity. These analyses evaluated whether neuroanatomical signals emerge from NeuroFM without task-specific adaptation.

NeuroFM was pre-trained exclusively on 100,000 AI-generated human brain volumes from the LDM100k dataset [28, 29], previously annotated with age, sex, total brain volume (TBV), and ventricular volume (Fig. 1; Method 2), allowing us to assess whether biologically relevant features can arise without exposure to real-world pathology. The model was subsequently evaluated on three deployment paradigms: direct prediction (predictor; Method 3), latent feature extraction, (encoder; Method 4), and dataset-specific fine-tuning (Method 5). These paradigms were used to test whether the learned representations capture biologically meaningful variations associated with clinical conditions or pathology.

**Fig. 1.**
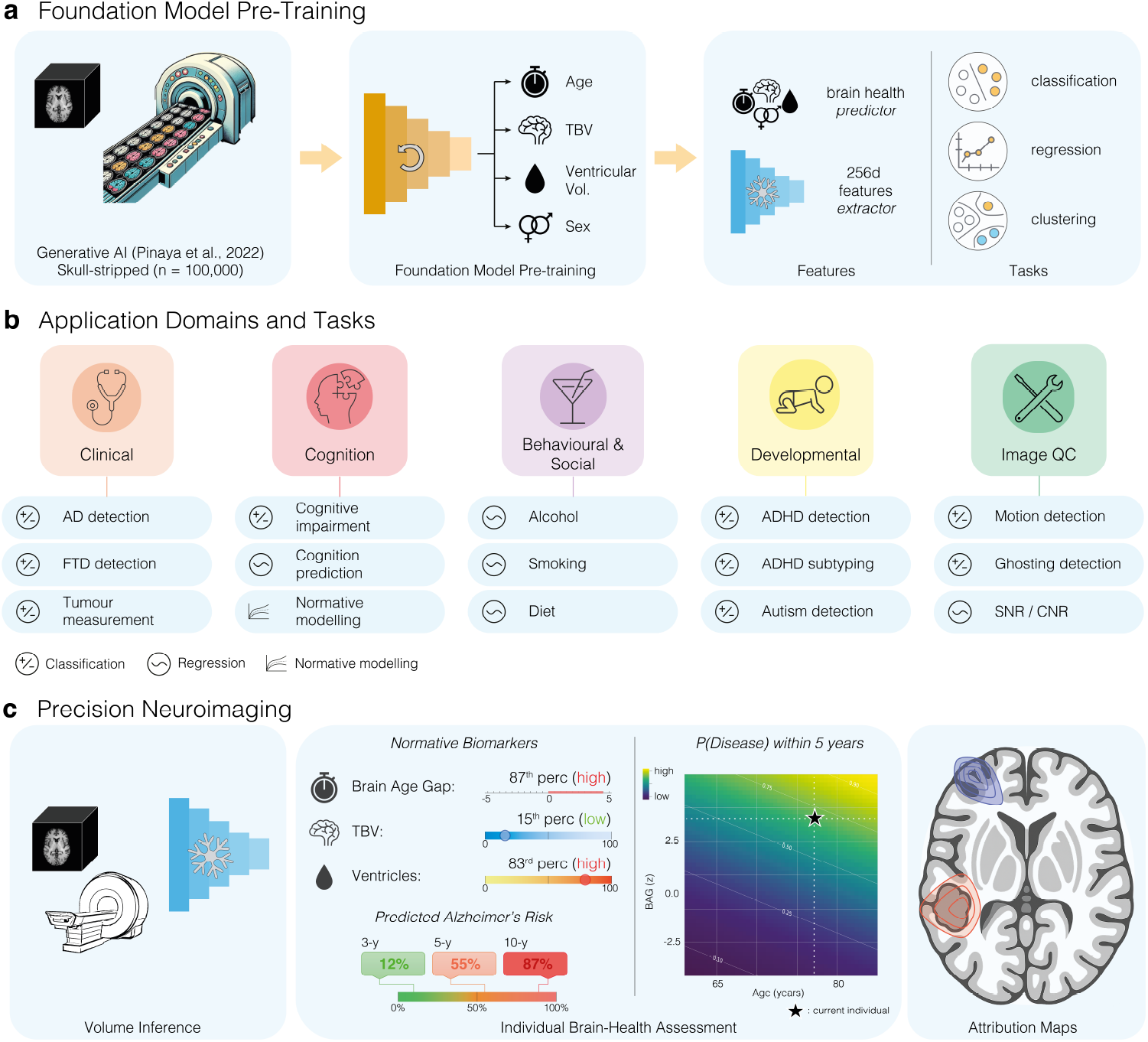
General overview. **a**, Foundation model (NeuroFM) pre-trained on 100,000 AI-generated volumes (left) using supervised learning targets for age, total brain volume (TBV), ventricular volume, and sex (middle). NeuroFM acts in two modes: predictor which gives four brain health estimates (age, sex, TBV, and ventricular volume), and encoder which extracts 100+ brain health features from each MRI. In this work, we apply NeuroFM to a variety of tasks, including classification, regression, and qualitative clustering analysis (right). **b**, After pre-training, the model is applied across five biologically motivated task subgroups: clinical, cognitive, socio-behavioural, developmental, and image quality metrics (left to right). **c**, By combining subtasks at inference (left), NeuroFM can create comprehensive, individualized health reports for a single individual (middle), with explainable attribution maps to highlight relevant brain regions (right).

Results are organized across five domains: clinical (Fig. 2), cognitive (Fig. 3), behavioural (Fig. 4), developmental (Fig. 5), and technical domains (Fig. 6). A final section examines whether these representations support true individualized brain-health estimation from a single structural MRI volume, including subject-level risk estimation and brain-health trajectories derived from single MRI volumes (Fig. 7), to progress from population-level modelling towards precision neuroimaging at the individual level.

**Fig. 2.**
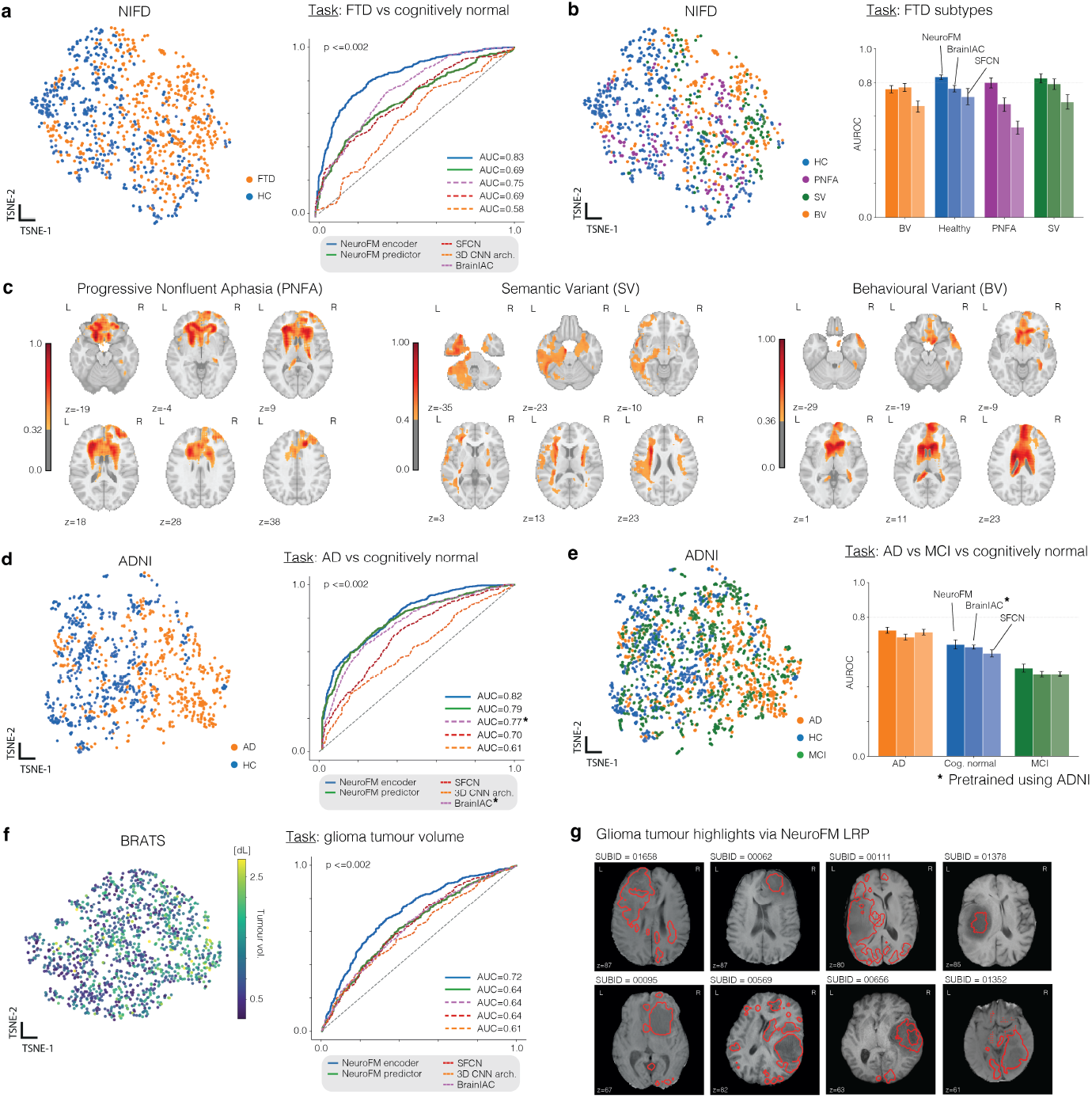
NeuroFM feature representations and diagnostic performance across datasets. For panels **a, b, d, e**, and **f**, the left subplot shows a t-SNE projection of NeuroFM extracted features coloured by class label, and the right subplot shows a comparison plot of classifier performance (expressed as AUC). **a**, Frontotemporal dementia (FTD, orange) versus cognitively healthy controls (HC, blue) in the NIFD dataset (Frontotemporal Lobar Degeneration Neuroimaging Initiative). NeuroFM features show group separation and achieve an AUC of 0.83 (p *≤* 0.002), outperforming the strongest competing method (AUC=0.75). **b**, FTD subtypes: primary non-fluent aphasia (PNFA, purple), behavioural variant FTD (BV, orange), semantic variant (SV, green), and healthy controls (blue). In one-vs-rest classification, NeuroFM achieves AUCs of PNFA = 0.80, BV = 0.76, SV = 0.83, and healthy = 0.83 (p *≤* 0.002). **c**, Layerwise Relevance Propagation (LRP) from NeuroFM predictions highlights anatomically relevant portions of the brain, related to FTD subtype (PNFA, SV, BV); coloured by relevance intensity. **d**, Alzheimer’s disease (AD, orange) versus cognitively healthy controls (HC, blue) on ADNI dataset (Alzheimer’s Disease Neuroimaging Initiative). AD vs HC prediction, NeuroFM compared to other models (NeuroFM encoder, AUC = 0.82, p *≤* 0.002). **e**, t-SNE projections of three-class ADNI classification (AD vs mild cognitive impairment (MCI, green) vs HC) show overlapping feature distributions across groups; one-vs-rest AUCs are AD = 0.73, MCI = 0.51, and healthy = 0.64 (p *≤* 0.002). **f**, BraTS-2023 glioma dataset [32]. Despite limited separation in extracted representation space (left, coloured by tumour volume), classification of large versus small tumours using encoder features achieves an AUC of 0.72 (p *≤* 0.002; right), with relevance maps localizing tumour regions. **g**, NeuroFM LRP maps for real glioma-affected brains shows distinct highlighting in focally affected brain regions (red).

**Fig. 3.**
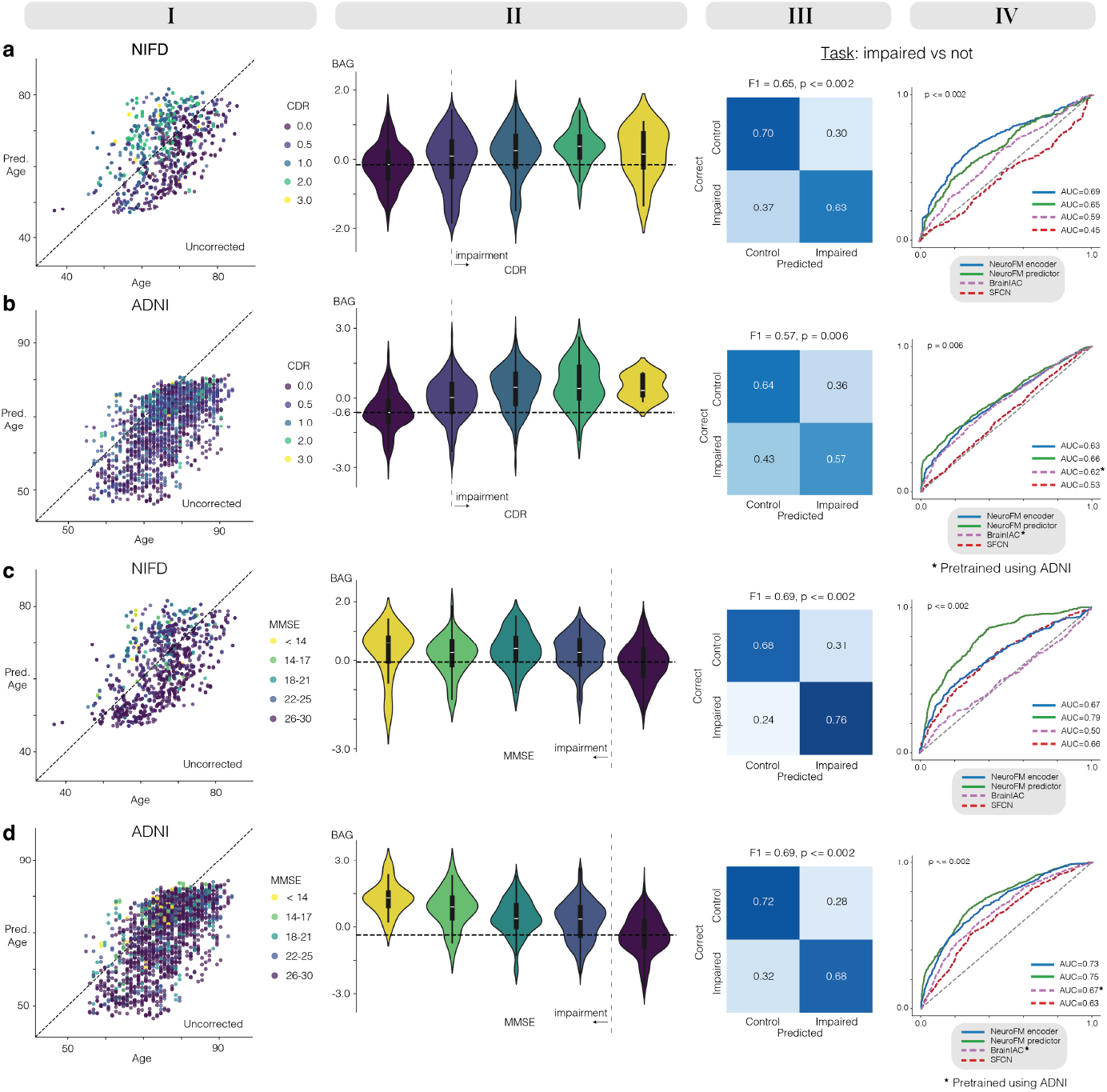
Associations between brain age gap and cognitive impairment in ADNI and NIFD. Predicted versus chronological age coloured by cognitive score (Column I); distribution of brain age gap (BAG) across levels of cognitive impairment (Column II); and binary classification of cognitively unimpaired versus impaired individuals using model-derived latent features (Columns III and IV). Classification performance is reported both as F1 scores and area under the receiver operating characteristic curve (AUC). **a**, Clinical Dementia Rating (CDR) in NIFD dataset. Individuals with CDR *>* 0 show a shift towards positive BAG relative to the unimpaired baseline. Impairment classification performance: F1 = 0.65 (p *≤* 0.002). **b**, CDR in ADNI dataset. BAG distributions are similarly shifted toward higher values in individuals with CDR *>* 0. Classification performance: F1 = 0.57 (p = 0.006). **c**, Mini-Mental State Examination (MMSE) in NIFD. MMSE scores below the impairment threshold are associated with positive BAG. Classification performance: F1 = 0.69 (p *≤* 0.002). **d**, MMSE in ADNI. Impaired individuals show higher mean BAG than cognitively unimpaired individuals. Classification performance: F1 = 0.69 (p *≤* 0.002).

**Fig. 4.**
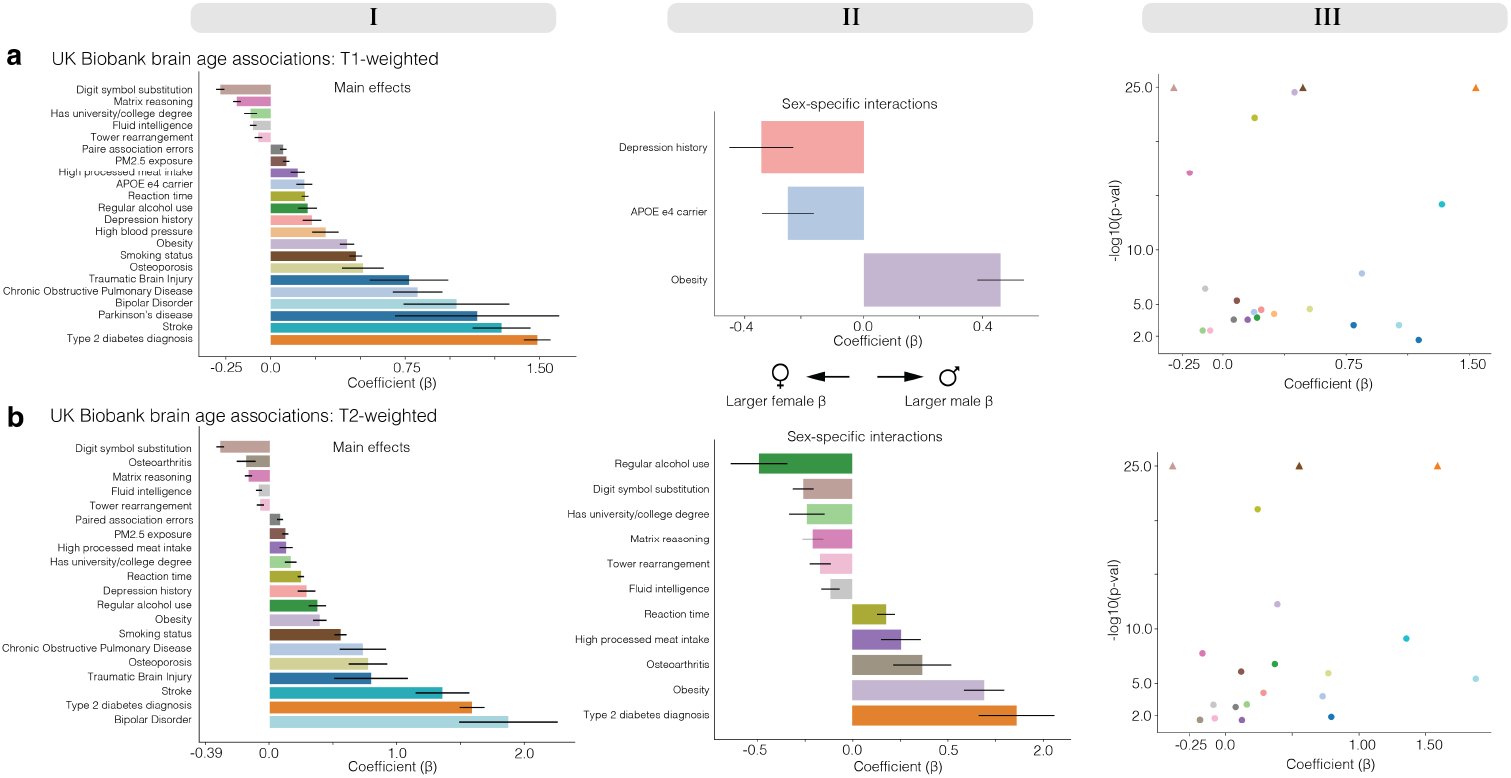
Clinical, lifestyle and cognitive associations with brain age gap and their moderation by sex. Models were fine-tuned on a subset of the UK Biobank (n = 7,700; 15,400 T1-weighted (T1w) and T2-weighted (T2w) volumes) and evaluated on the held-out remainder (n = 36,943; 82,904 T1w and T2w volumes). T1w and T2w input sequences were analysed separately. All coefficients were corrected for multiple comparisons using the Benjamini–Hochberg false discovery rate (FDR) method. Column I: standardized regression coefficients (*β ±* s.e.m.) for main effects on brain age gap (BAG; predicted minus chronological age). Column II: sex-interaction coefficients (*β ±* s.e.m.) for variables showing significant moderation by sex. Column III: volcano plots displaying effect size (x axis) against statistical significance (y axis) for main effects; capped values are denoted by triangles. **a**, T1w MRI analysis. Higher BAG (older-appearing brain relative to chronological age) most strongly associated with history of type 2 diabetes, stroke, Parkinson’s disease, bipolar disorder and traumatic brain injury. Lower BAG was associated with higher cognitive performance and greater educational attainment. Sex-interaction analyses revealed stronger BAG associations in males for obesity and processed meat intake, and in females for history of depression. **b**, T2w MRI analysis. Main effects were broadly consistent with the T1w analysis, although the effect of bipolar disorder differed in magnitude and the association with Parkinson’s disease was no longer significant. The most prominent divergence from the T1w results appeared in sex-specific interactions: BAG associations were stronger in males for type 2 diabetes and osteoarthritis, and in females for regular alcohol use.

**Fig. 5.**
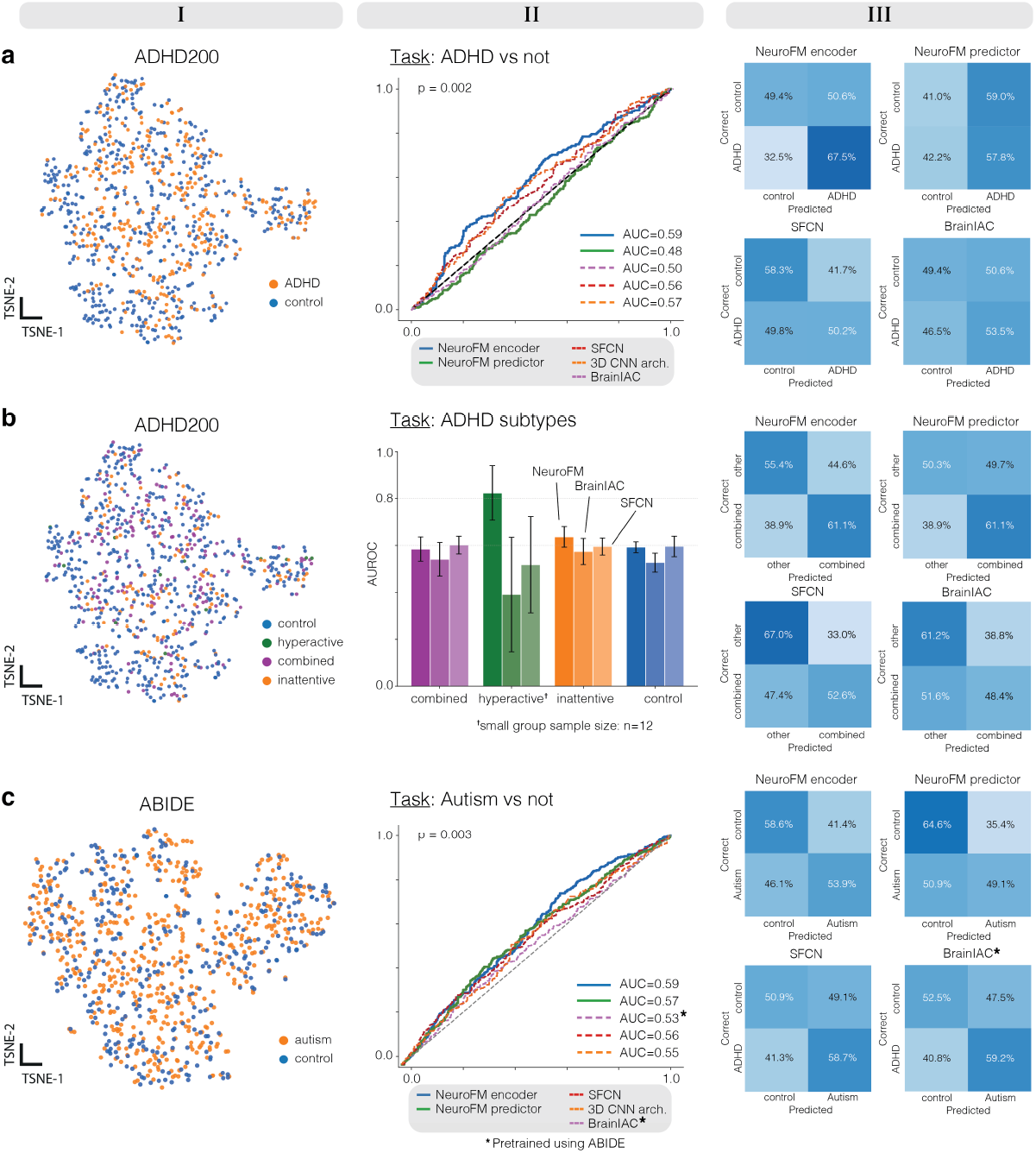
Neurodevelopmental analyses across ADHD and autism datasets. t-SNE projections of NeuroFM extracted features (Column I); diagnostic classification performance using NeuroFM features expressed as area under the receiver operating characteristic curve (AUC) (Column II); and corresponding confusion matrices for model predictions (Column III). **a**, ADHD200 dataset [52], attention-deficit/hyperactivity disorder (ADHD) versus controls. t-SNE projections do not show clear separation between ADHD-positive (orange) and control (blue) individuals. Even so, classification using NeuroFM extracted features achieves an AUC of 0.59 (p = 0.002). **b**, ADHD200 subtype differentiation across control (blue), hyperactive (green), combined (purple), and inattentive subtypes (orange). t-SNE projections show overlapping feature distributions across subtypes. One-vs-rest classification yields NeuroFM AUCs of non-ADHD = 0.59, hyperactive = 0.82, combined = 0.58, inattentive = 0.64. **c**, ABIDE dataset [53, 54], autism spectrum disorder (orange) versus controls (blue). t-SNE projections show no clear diagnostic clustering. Classification using NeuroFM extracted features achieves an AUC of 0.59 (p = 0.003).

**Fig. 6.**
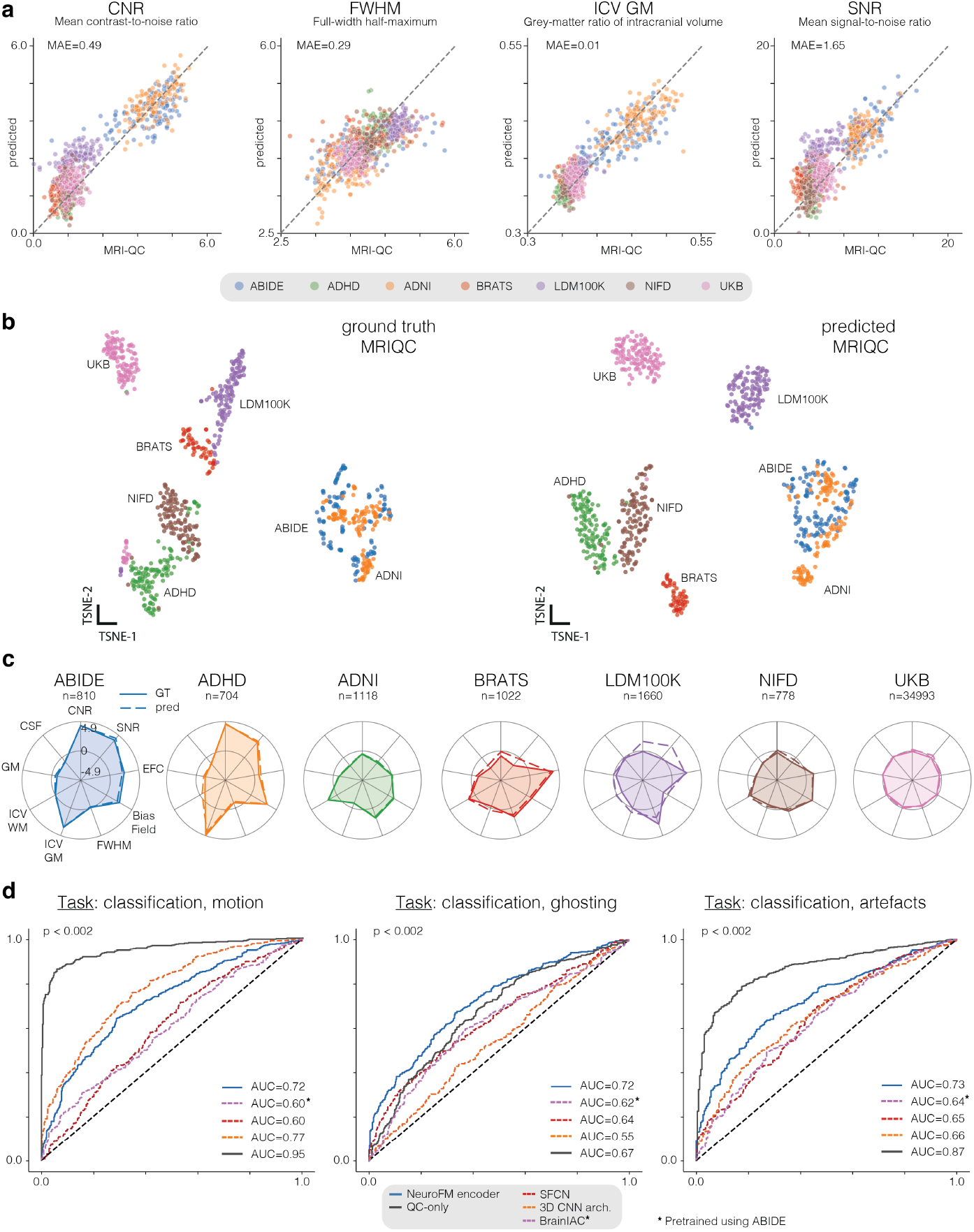
Image quality and artefact identification. **a**, Scatter plots comparing NeuroFM-predicted vs ground-truth MRIQC [55] metrics across test volumes from seven datasets. Predictions were generated by fitting linear regressors to frozen NeuroFM features (Method 4). Mean prediction nMAE (normalized MAE) across all test datasets was 1.24 (std. 0.6). Definitions for all metrics are provided in Supplementary Table ST2. **b**, Two-dimensional t-SNE projections of ground-truth MRIQC metrics (left) and NeuroFM predictions (right), coloured by dataset. Comparable within-cohort clustering indicates that predicted metrics preserve dataset-level structure in QC feature space. **c**, Radial plots summarizing nine z-scored QC metrics for each dataset. Solid lines denote ground-truth means and dashed lines denote NeuroFM predictions, illustrating close agreement in cohort-specific artefact profiles. **d**, AUROC curves for classification of synthetic motion, ghosting, and combined artefacts. Models trained on MRIQC metrics showed the highest accuracy for motion-related artefacts, whereas NeuroFM features performed best for ghosting, demonstrating complementary detection characteristics.

**Fig. 7.**
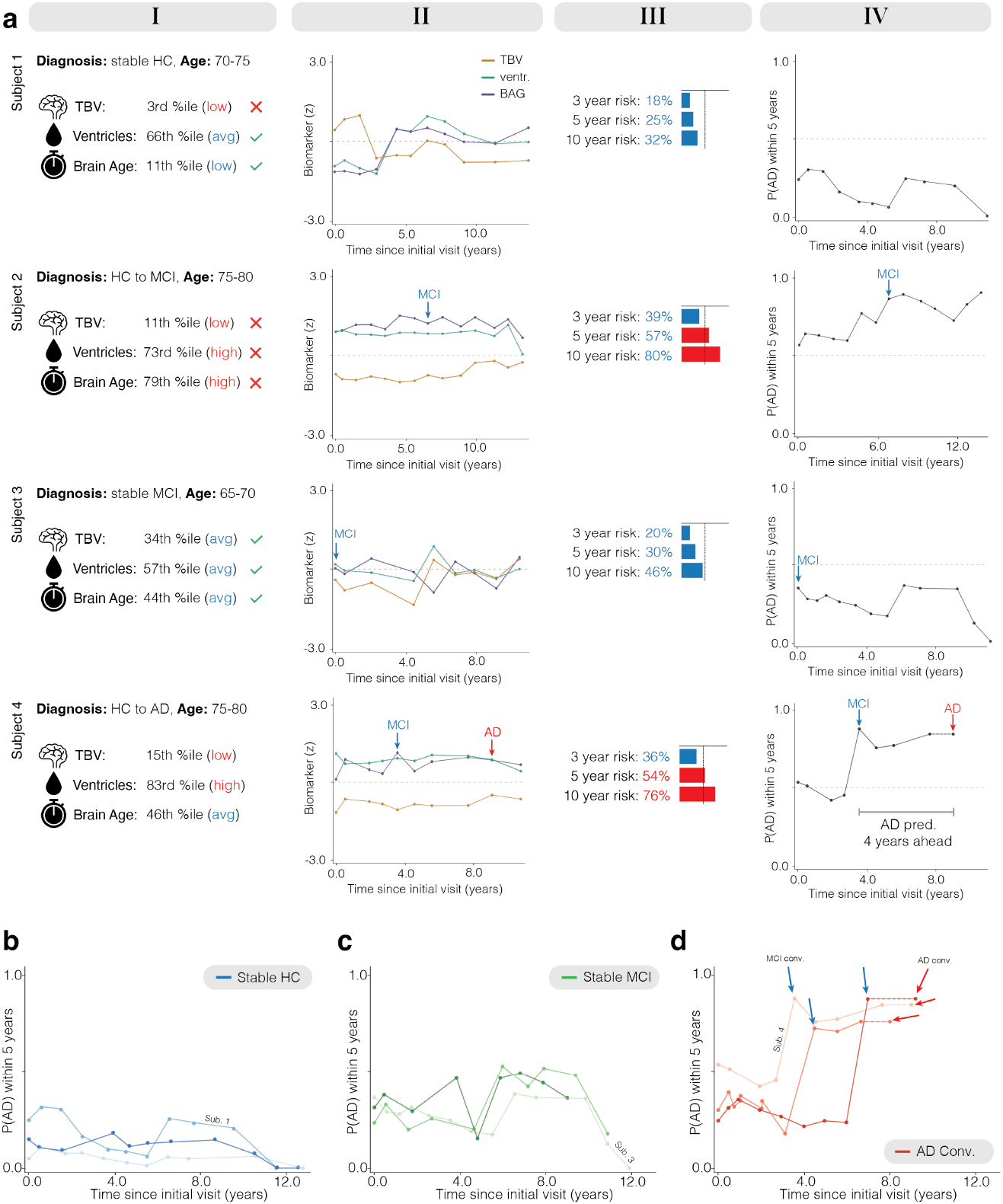
Precision neuroimaging enables individualized prediction of Alzheimer’s disease risk. Individual-level longitudinal trajectories are shown for nine representative individuals from the Alzheimer’s Disease Neuroimaging Initiative (ADNI) cohort. Normative models were constructed using NeuroFM predictor outputs (Methods 3 and 10), on which Cox hazards model were applied to estimate longitudinal risk of Alzheimer’s dementia (Method 11). Time is reported in years since the initial study visit. **a**, (Column I) Baseline normative estimates of characteristics (total brain volume (TBV); ventricular volume; and brain age gap (BAG)), where were derived at first visit for each individual and remained relatively stable over time. (Column II) Normative feature estimates across visits, with diagnostic events indicated. (Column III) Predicted baseline probabilities of AD diagnosis at 3, 5, and 10 years calculated at baseline. Red indicates the estimated risk of conversion is over 50%. (Column IV) Rolling 5-year predicted probability across visits, with diagnostic events highlighted. **b**, Individuals who remained cognitively normal over approximately 12 years maintained predicted 5-year probability under 50%. **c**, Individuals diagnosed with mild cognitive impairment (MCI) at baseline who did not progress to AD also showed predicted 5-year risk below 50%, although estimates were generally higher than in cognitively normal individuals (panel **b**). **d**, Individuals who progressed to AD exhibited increasing predicted 5-year risk over time, with the steepest rise occurring after MCI diagnosis.

### 2.1 Clinical pathology classification without task-specific adaptation

NeuroFM encodes biologically meaningful representations of brain health that generalize across pathological and imaging contexts that were unseen during pretraining. We evaluated its representational encoding across three progressively stringent settings: detection of neurodegenerative disease, differentiation of clinically overlapping subtypes, and characterization of focal tumour pathology in an out-of-domain dataset. To isolate biological signal from acquisition-related confounds, all analyses were performed within a confound-robust framework, which treats MRI quality metrics as nuisance variables using double machine learning (Method 7).

Neurodegeneration is a demanding test because morphometric signatures are spatially distributed and often subtle; we therefore assessed detection and differentiation of both Alzheimer’s disease (AD) and frontotemporal dementia (FTD; Method 6.a).

Gliomas provide a complementary challenge through focal rather than distributed structural disruption (Method 6.b). In both settings, features were extracted from frozen encoders and evaluated using logistic regression.

#### Frontotemporal dementia

Using the Neuroimaging in Frontotemporal Dementia (NIFD) dataset (n = 259 individuals; 1,451 MRI volumes), NeuroFM reliably distinguished FTD patients from controls (AUC = 0.83; p ≤ 0.002; Fig. 2a), outperforming a ResNet-style CNN (AUC=0.58) [33], a pre-trained Simple Fully Convolutional Network (SFCN; AUC=0.69) [19], and Brain Imaging Adaptive Core (BrainIAC; AUC=0.75; Method 13) [22]. NeuroFM subtype prediction yielded AUCs of 0.76-0.83 (p ≤ 0.002; Fig. 2b). Unsupervised inspection of extracted features with t-SNE revealed spontaneous separation of healthy and diseased individuals, with partial separation of FTD subtypes (Fig. 2a,b (left); Method 3). Relevance maps (Fig. 2c; Method 8) aligned with established neuroanatomical profiles: nonfluent variant (PNFA) relevance concentrated in left inferior frontal cortex and anterior insula [34]; semantic variant in left temporal pole, amygdala, and hippocampus [35]; and behavioural variant in anterior cingulate, medial prefrontal cortex, and striatum [36]. **Alzheimer’s disease**. Using the Alzheimer’s Disease Neuroimaging Initiative (ADNI) dataset (n=265 individuals; 2,450 MRI volumes), NeuroFM achieved AUC = 0.77 (p ≤ 0.002) for AD versus cognitively health classification (HC), and AUCs of 0.51-0.73 for AD vs MCI vs healthy comparisons (p ≤ 0.002; Fig. 2d,e). NeuroFM’s lower differentiation performance relative to FTD is consistent with the more subtle and heterogeneous morphological presentation of AD [37] and was obtained without exposure to pathological volumes during pretraining, with per-class performance comparable to prior work [19, 22]. Notably, NeuroFM outperformed BrainIAC despite the latter being pretrained on ADNI data that potentially included scans in our evaluation. **Glioma characterization**. In the Brain Tumor Segmentation 2023 (BraTS-2023) dataset (n=1,127 individuals; 1,244 volumes) [32], tumour masks defined a binary volume classification task relative to the dataset median (Fig. 2f; Method 6.b), yielding significant NeuroFM performance after artefact correction (AUC = 0.72; p *<* 0.002). LRP maps consistently localized tumour regions within the source MRI (Fig. 2g), confirming sensitivity to focal pathology.

Together, these results demonstrate that NeuroFM captures biologically meaningful structural variation that generalizes across both distributed and focal pathologies, despite being trained exclusively on healthy synthetic data. These findings support the use of anatomically grounded representations for cross-domain, disease-agnostic inference from structural MRI.

### 2.2 Cognition impairment and brain age gap

We next asked whether NeuroFM representations capture variation in cognitive function, a continuous phenotype that lacks a discrete anatomical boundary and is thus more challenging to recover from structural MRI than categorical diagnosis. Whereas diagnostic classification exploits group-level anatomical differences, cognitive decoding requires the representation to track distributed variation in brain health (Method 6.c). **Cognitive status is reliably decodable from the latent space**. Linear classifiers trained on frozen NeuroFM representations discriminated impaired from normal cognition above all baselines (Fig. 3, cols. III-IV; Methods 13). In NIFD, AUC for CDF reached 0.69 (NeuroFM) vs 0.59 (BrainIAC; p ≤ 0.002) and for MMSE, 0.78 (NeuroFM) vs 0.66 (SFCN; p ≤ 0.002). In ADNI, AUC for CDR reached 0.66 (NeuroFM) vs 0.62 (BrainIAC) and for MMSE, 0.75 (NeuroFM) 0.75 vs 0.67 (BrainIAC; both p = 0.006). Confusion matrices indicated balanced sensitivity and specificity across both tasks.

#### Brain age gap is associated with cognitive impairment

To provide an interpretable summary of this relationship, we examined the brain age gap (BAG), which can be viewed as a one-dimensional projection of the underlying multidimensional representation of brain health. BAG is defined as the difference between the predicted and chronological brain age. It is a biomarker which is informative across a range of clinical, social, and behavioural outcomes, with positive values indicating accelerated brain ageing [18, 21, 38–41]. All analyses were adjusted for age and acquisition covariates to isolate cognition-associated effects (Method 7; see Supplementary Fig. S14 for unadjusted association).

BAG was significantly associated with cognitive function across both cohorts and measures (Fig. 3, col. I-II). In the NIFD dataset (n=259; 1,115 volumes), BAG increased with worsening CDR (*β* = 0.26; p *<* 0.0001) and with lower MMSE scores (*β* = -0.23; p *<* 0.0001). Associations were comparable in ADNI (n=265; 2,408 volumes) for CDR (*β* = 0.33; p *<* 0.0001) and MMSE (*β* = -0.37; p *<* 0.0001).

### 2.3 Socio-behavioural factors of brain ageing

To evaluate the population-level contributors of brain health captured by NeuroFM, we fine-tuned the model on a healthy subset of UK Biobank individuals (n = 7,700; 15,400 T1w and T2w volumes; UK Biobank project 17689) [42] and quantified associations between clinical, cognitive and lifestyle variables and the brain age gap (BAG; Method 9). Fine-tuning, rather than linear regression on frozen features, was performed separately for T1w and T2w sequences to enable adaptation to a modality not seen during pretraining; associations from the frozen model are reported in Supplementary Fig. S15. BAG associations were tested on held-out participants (n = 36,943; 82,804 T1w and T2w volumes) using fully adjusted linear models controlling for age and sex.

#### Brain age gap reflects established health and cognitive correlates

Greater BAG was associated with prevalent type 2 diabetes, stroke, Parkinson’s disease, bipolar disorder, and traumatic brain injury (all p *<* 0.001 after FDR correction; Fig. 4, col. I), while higher cognitive performance and educational attainment (university and college degree versus not) were associated with lower BAG. Main effects were broadly consistent across T1w and T2w analyses, with two notable exceptions; Parkinson’s disease association was attenuated in T2w, and the bipolar disorder effect differed in magnitude. This plausibly reflects the distinct tissue properties each sequence captures. Moreover, T2w contrast is additionally sensitive to white matter water content, periventricular changes, and brain iron [43]. These patterns align with prior large-scale BAG studies [38, 44], confirming that NeuroFM recovers known population-level signatures of brain ageing from pre-trained model inference (Supplementary Fig. S15) and after fine-tuning to a new cohort and modality (Fig. 4).

#### Sex-dependent variation in brain health associations

Interaction models identified BAG associations differed significantly between males and females across several exposures (all interaction p *<* 0.05 after FDR correction; Fig 4, col II). In T1w analyses, a larger BAG interaction coefficient associated with depression history and *APOE ϵ*4 carrier status in females, and with obesity in males. T2w analyses revealed a partially distinct profile: a larger BAG interaction coefficient for regular alcohol use in females, and for type 2 diabetes and osteoarthritis in males. The female-specific alcohol effect, emerging selectively in T2w, might reflect T2-specific tissue changes associated with alcohol consumption, including white matter disruption [45] and subcortical iron accumulation [46], though whether these mechanisms underlie the observed sex difference warrants further investigation. The divergence in sex-interaction profiles across modalities demonstrates that fine-tuning NeuroFM to T2w surfaces biologically distinct signatures, including patterns consistent with sex-specific vulnerability in neurodegeneration [47, 48], that would remain hidden in single-sequence analyses.

### 2.4 Neurodevelopmental disorder analyses

Having established performance in disorders with pronounced structural signatures, we next evaluated NeuroFM in conditions where anatomical differences are subtle, heterogenous, and inconsistently reported. Neurodevelopmental disorders such as attention-deficit/hyperactivity disorder (ADHD) and autism provide a stringent lower-bound test of representational sensitivity [49–51]. NeuroFM was evaluated on the ADHD-200 dataset (n = 880) [52] and the Autism Brain Imaging Data Exchange dataset (ABIDE; n = 904; Method 6.d) [53, 54]. Extracted features showed weak but consistent differentiation across ADHD, ADHD subtypes, and autism (Fig. 5), with t-SNE projections revealing no clear diagnosis-specific clustering, arguing against separability driven by coarse morphometric variation such as total brain volume or ventricle size.

#### Detection of subtle neurodevelopmental signals

ADHD classification by NeuroFM reached AUC = 0.59 (p = 0.002; Fig. 5a), modestly exceeding the strongest obtained baseline (AUC = 0.57, 3D CNN arch.), and yielding above-chance subtype differentiation for hyperactive and inattentive subtypes. Reliable performance was observed only for the large NeuroFM encoder variant (NeuroFM-L), while smaller models did not exceed chance levels (Supplementary Figs. S6-S7), indicating that higher representational capacity is required to capture the weak, potentially nonlinear structural effects. The morphometric baseline, comprised of NeuroFM-derived estimates of age, sex, total brain volume, and ventricular volume, performed worse than the full embedding, indicating that the representation encodes variation beyond global anatomy. In ABIDE, NeuroFM autism classification reached AUC = 0.59 (p = 0.003), again slightly exceeding all baselines. Notably, this included BrainIAC, which was pretrained on both ABIDE and ABCD, yet was still outperformed by NeuroFM pretrained exclusively on synthetic healthy adult data. Confusion matrices were not markedly asymmetrical (Fig. 5c, col. III), indicating that class imbalance does not explain the observed effects.

#### Interpretation within a low-effect regime

Small effect sizes are expected given the limited and heterogeneous structural findings reported for ADHD [50, 51] and autism [49]. NeuroFM embeddings showed substantial group overlap after adjustment for age and quality-control confounds (Fig. 5, col. I; Method 7). Nonetheless, above-chance performance across both datasets indicates that the encoded NeuroFM representation captures reproducible, albeit weak, disorder-related signal without relying on coarse morphometric proxies. Comparable baseline performance suggests that the overall classification performance ceiling reflects limited structural signal available in MRI for these conditions rather than a limitation of any particular model.

### 2.5 Disentangling biological associations from acquisition-related variation

Reliable person-level inference requires representations that distinguish biological variation from acquisition-related artefacts [24]. We evaluated whether NeuroFM encodes image-quality structure consistent with established quality control (QC) metrics while remaining robust to heterogeneous scanner conditions (Supplementary Figs. S3, S17).

Because template-based QC pipelines frequently fail when volumes cannot be reliably registered to standard space (Supplementary Table ST1), this analysis also assesses NeuroFM as a template-free alternative for image-quality assessment.

#### Recovery of established quality metrics

Linear regression models trained on NeuroFM features predicted commonly used MRIQC metrics (Methods 4, 12.a) [55] across multiple cohorts (Supplementary Table ST4). Reliable QC prediction was observed for the small encoder variant, whereas larger models did not perform reliably on this auxiliary task under linear probing (Supplementary Figs. S1, S3; Method 12). M and L variants nevertheless showed lower QC confounding than did comparison models prior to residualization (Supplementary Fig. S8). This finding indicates that larger encoders increasingly disentangle acquisition from biological variation during pretraining rather than requiring post-hoc correction. Across held-out volumes, predictions showed strong agreement with MRIQC outputs in under 10 seconds per volume without template-related failures. Error was lowest for contrast-to-noise ratio (CNR; MAE = 0.49), and for full-width at half maximum (FWHM; MAE = 0.29), and signal-to-noise ratio (SNR; MAE = 1.65; Fig. 6a-c; Supplementary Fig. S3), indicating that acquisition-related variation is explicitly encoded within the feature space rather than implicitly driving downstream predictions.

#### Direct detection of imaging artefacts

We tested whether artefacts were directly decodable from NeuroFM features by adding motion and ghosting augmentations to ABIDE scans (Method 12.b), comparing linear classifiers against MRIQC-derived features and baseline models (Methods 12.b, 13). MRIQC features performed best for motion (AUC = 0.95 vs 0.72), and combined motion plus ghosting (AUC = 0.87 vs 0.72), whereas NeuroFM performed best for ghosting alone (AUC = 0.72 vs 0.67), outperforming both SFCN [19] and BrainIAC [22] across all artefact conditions. This indicates NeuroFM is sensitive to structured replication artefacts less fully captured by conventional QC metrics (Fig. 6d). Together, these findings suggest that acquisition-related variation is measurable within the representation yet does not account for the observed biological associations.

### 2.6 Individual risk trajectories

A central objective of precision neuroimaging is to generate prospective, person-level estimates from a single structural MRI. We therefore evaluated whether NeuroFM-derived representations support individualized forecasting of future dementia risk without task-specific adaptation. Using normative trajectories derived from cognitively normal ADNI individuals (n=1,594; 10,893 volumes), elevated BAG was associated with increased risk of conversion to Alzheimer’s disease, detectable up to five years before diagnosis via Cox proportional hazards models (Methods 10-11; Fig. 7d; Extended Data Fig. 2). ADNI data were used solely to fit downstream normative and survival models; the NeuroFM encoder was fixed throughout without finetuning or task adaptation.

#### Population-derived models capture prospective dementia risk

At the population level, estimated AD risk increased monotonically with age and BAG, supporting survival-model calibration (Extended Data Fig. 2b-e). Linear mixed-effects analyses, controlling for age and sex, revealed a significant main effect of BAG group on outcome trajectory in the 5 years preceding conversion (*β* = 1.844, p *<* 0.002), with those converting from MCI to AD averaging 1.52 years higher than non-converters over this period (Method 14; Extended Data Fig. 2a). Although these analyses establish population-level validity rather than individualized prediction, their convergence strengthens confidence in the underlying risk signal.

#### Foundation-model representations enable individualized risk trajectories

We applied these population-derived models to held-out individuals to test whether they produce clinically interpretable longitudinal forecasts. Using a fixed Cox proportional hazards model (Method 11) [56], we evaluated individuals, who were excluded from model fitting, across repeated visits. Longitudinal risk trajectories remained low among non-progressors, intermediate in stable MCI cases, and increased preceding diagnosis among converters (Fig. 7; Extended Data Fig. 2f). Notably, this divergence emerged despite relatively stable morphometric estimates (Fig. 7a, col. II), indicating that risk reflects persistent deviation from age-expected norms rather than rapid structural change.

## 3 Discussion

Our study demonstrates that a single structural MRI foundation model can capture a broad spectrum of information relevant to brain structure and brain health. Trained once on a large corpus of artificially generated MRIs using proxy morphometric and demographic targets, NeuroFM learns representations that generalize across tasks, datasets, and domains without task-specific optimization [57]. Across independent cohorts, simple linear models trained on NeuroFM features accurately predicted neurodegenerative disease status (Fig. 2a-d), cognitive performance (Fig. 3), socio-behavioural associations (Fig. 4), neurodevelopmental disorders (Fig. 5), tumour characteristics (Fig. 2e), and image quality assessment (Fig. 6), and, crucially, individual risk trajectories (Fig. 7). The consistent performance of these representations indicates that NeuroFM functions as a reusable representation that preserves biologically meaningful information about brain anatomy, rather than as a task-specific classifier.

### Precision neuroimaging as a paradigm shift

Achieving precision neuroimaging requires moving beyond task-specific models toward population-scale representations of brain anatomy that enable person-level inference. NeuroFM advances this paradigm by organizing structural MRIs into reusable population-level dimensions of anatomical variation. This paradigm shift directly addresses long-standing challenges in neuroimaging, including limited sample sizes, the impracticality of acquiring multiple data points per individual [58, 59], site variability [9], restricted generalizability, and poor comparability across studies [60]. Crucially, embedding individual scans within normative trajectories derived from large cohorts enables individualized inference from a single MRI volume (Fig. 7). In doing so, NeuroFM moves structural MRI closer to a scalable framework for quantifying brain health across individuals and populations.

### Brain age as a single dimension of multidimensional brain health

A key implication of this approach is that commonly used scalar biomarkers, such as the brain age gap (BAG), can be seen as being projections of a broader, multidimensional representation of brain health [61]. In both NIFD and ADNI, the NeuroFM-derived BAG showed robust associations with cognitive impairment, increasing with worsening CDR and lower MMSE scores (Fig. 3), consistent with prior brain-age literature [38]. However, the same latent NeuroFM representation also enabled the direct prediction of diagnostic labels for frontotemporal dementia and Alzheimer’s disease (Fig. 2), outperforming scalar morphometric baselines despite never being exposed to pathological volumes during pretraining. Importantly, results showed that diagnostic information is encoded across cortical and subcortical regions, rather than being driven solely by global volumetric measures or by a single volumetric feature (i.e., hippocampus volume). These findings demonstrate that disease-specific structural patterns emerged naturally from the representation and that BAG deviations reflect projections of this broader anatomical space [62, 63]. Consistently, NeuroFM enabled tumour size classification and localized tumour-relevant regions in the brain tumour dataset, BraTS (Fig. 2e), despite being pretrained exclusively on healthy, synthetic data, highlighting the representation’s ability to capture broadly useful structural priors [26].

### NeuroFM embeddings enable task-specific inference

NeuroFM revealed meaningful associations between brain structure and socio-behavioural factors in the UK Biobank dataset. After finetuning, NeuroFM BAG estimates were associated with metabolic, neurological, and psychiatric conditions, as well as cognitive performance and educational attainment (Fig. 4), consistent with existing literature [44]. Sex-specific interaction effects for variables such as depression, obesity, and *APOE ϵ*4 status suggest that the model captures both shared and stratified risk patterns. Associations emerged across multiple imaging modalities (T1w and T2w), indicating that NeuroFM encodes general structural correlates of brain health rather than task-specific signals.

In contrast, neurodevelopmental disorder classification highlighted the limits of structural MRI-based precision (Fig. 5). In ADHD and autism, NeuroFM embeddings achieved modest but reproducible classification performance, with overlap between diagnostic groups. This reflects the known heterogeneity and subtlety of structural effects in these conditions, indicating that even large-scale representations cannot recover strong diagnostic signals when the underlying anatomical differences are weak or diffuse. Nevertheless, performance exceeded baseline models, suggesting that NeuroFM captures structural variation beyond age, sex, and brain size. Beyond disease and cognition, NeuroFM embeddings also supported prediction of MRI quality metrics across multiple datasets (Fig. 6). The ability to approximate multiple QC measures from a shared representation suggests that structural priors learned for brain-health estimation also encode, and potentially disentangle, information about image quality and artefacts, enabling faster and more robust quality-control pipelines.

### Precision neuroimaging and understanding individual risk

NeuroFM enables precision neuroimaging through individualized risk estimation anchored to population-derived normative trajectories. By combining NeuroFM-derived normative models with survival analysis, deviations in predicted brain-health measures were translated into individual estimates of future dementia risk years before clinical diagnosis (Fig. 7) without requiring repeated within-individual measurements or task-specific models [26, 58, 64]. Representative trajectories illustrate how NeuroFM identifies divergence from expected trajectories, detecting elevated future risk in cognitively normal individuals, as well as stable trajectories in those with baseline impairment, going beyond diagnostic labels. Remarkably, these risk estimates arise from population-trained structural representations learned without exposure to Alzheimer’s disease-specific imaging data. This capability may support the development of standardized quantitative brain-health reporting, early risk stratification in research and clinical settings, and longitudinal monitoring. Consistent with this interpretation, adapting NeuroFM to a dense, single-subject longitudinal dataset collected across imaging sites (73 scans) [65, 66] demonstrates that the representation can be specialized to capture person-specific anatomical trajectories (Supplementary Fig. S17).

### Synthetic MRI enables cohort-agnostic brain health modelling

Prediction models trained on real-world neuroimaging data are vulnerable to multiple sources of confounding, including scanner-specific acquisition features, demographic biases, and co-occurring pathologies that can be exploited as predictive shortcuts rather than reflecting true neurobiological signal [24, 67, 68]. Synthetic imaging data generated from models trained on healthy populations offers a principled alternative for learning population-level anatomical representations with reduced confounding. By sampling from a generative distribution, synthetic data reduces the coupling between biological targets and incidental correlates while enabling unbounded labelled datasets, bypassing privacy constraints. Importantly, synthetic MRI volumes are designed to resemble real, in-vivo human data while avoiding biases introduced by condition-specific alterations [68], supporting their generalization to clinical cohorts. Additionally, unlike multi-cohort real-world datasets, the LDM100k dataset [28, 29] provides a uniform age distribution that reduces prediction bias in distribution tails compared with existing models (Supplementary Fig. S18), while preserving accurate morphometric estimation on unseen external cohorts (Supplementary Fig. S10). Consistent with this, NeuroFM exhibits reduced confound sensitivity compared with models trained with real-world data (Supplementary Figs. S6, S7, S8). To further mitigate residual confounds, we applied double machine learning to remove residual linear confound relationships (Method 7).

### Limitations and future work

Several factors delimit the scope of this work.

Pretraining on synthetic structural MRI enables scale and diversity but may introduce biases that differ from real-world clinical data. The framework is cross-sectional and does not model longitudinal trajectories or disease progression. Moreover, NeuroFM operates exclusively on structural MRI and does not incorporate functional, molecular, or extracerebral factors central to broader conceptions of brain health [69]. NeuroFM should therefore be interpreted as providing a foundational structural representation, rather than a comprehensive or mechanistic model of brain disorders. Although NeuroFM supports spatial attribution maps that consistently highlight anatomically plausible regions across tasks, interpretability remains limited. These maps should not be interpreted as mechanistic explanations or causal evidence, but rather as tools that provide transparency at the individual level.

Future work should evaluate prospective performance in independent clinical cohorts, particularly using longitudinal designs to test stability and forecasting values. Extending the framework to integrate multimodal imaging and non-imaging biomarkers may clarify how structural representations relate to broader dimensions of brain health. Further work is also needed to characterize how acquisition-related variation and biological signal coexist within foundation model representations.

To conclude, NeuroFM reframes precision neuroimaging as a scalable foundation-model paradigm for individualized brain-health assessment from routine structural MRI.

## 4 Methods

### 4.1 Method 1: Data and preprocessing

#### Datasets

We aggregated 18 distinct publicly accessible datasets, comprising 236,355 unique MRI volumes from 161,757 individuals, spanning healthy populations and a broad range of neurological and psychiatric conditions (Fig. 8). These included UK Biobank (n=44,790, 98,598 volumes; Sudlow et al., 2015), ADNI (n=2,011; 17,320 volumes; Alzheimer’s Disease Neuroimaging Initiative), NIFD (n=346; 1,847 volumes; Frontotemporal Lobar Degeneration Neuroimaging Initiative), IXI (n=569, 569 volumes), a publicly available structural MRI dataset, OASIS-3 (n=1,025; 2,633 volumes) [70], LDM100k (n=99,994; 99,994 volumes) [28, 29], ADHD200 (n=880; 880 volumes) [52], ABIDE (n=904; 904 volumes) [53, 54], FCON1000 (n=6,904; 7,820 volumes) [71], AOMIC (n=931; 1,911 volumes) [72], BraTS 2023 (n=1,127; 1,244 volumes) [32], EDSD (n=288; 290 volumes) [73], HCP (n=890) [74], IBSR (n=18 [RRID:SCR_001994](http://www.nitrc.org/projects/ibsr)), MindBoggle101 (n=92) [75], MRBrainS (n=7) [76], and SIMON (n=1; 73 volumes) [65, 66]. Only the LDM100k dataset was used for training the baseline foundation model. All the remaining datasets were reserved exclusively for downstream evaluation and task-adaptation experiments. Demographic coverage and age/sex distributions across datasets are reported in Fig. 8. All reported volume and individual counts reflect totals used across the study and may vary by analysis depending on the availability of diagnostic or cognitive data; exact sample sizes for each task are provided in Supplementary Tables ST1, ST3, and ST4.

**Fig. 8.**
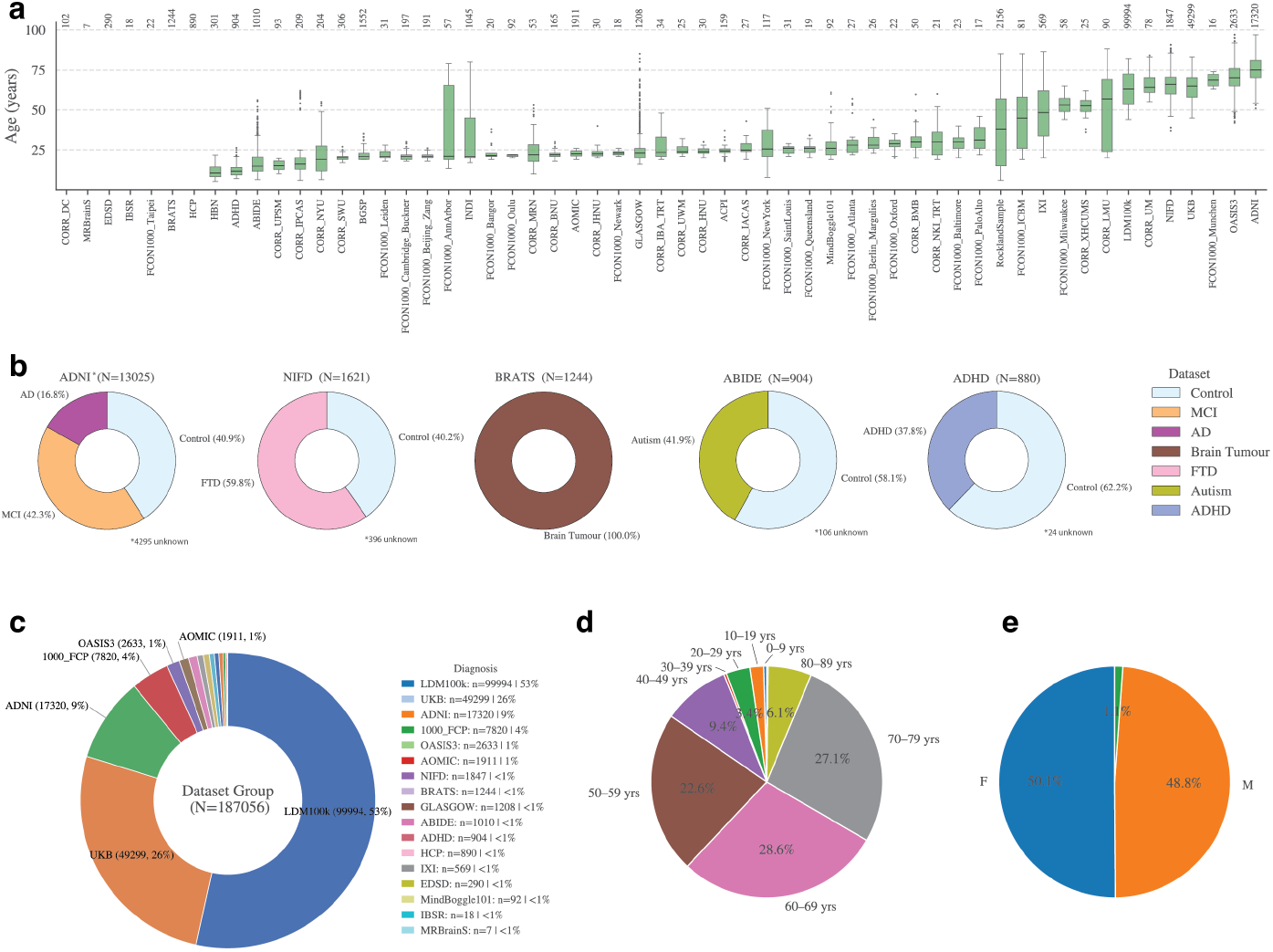
Overview of datasets used for NeuroFM training and evaluation. **a**, Age distributions across all included datasets, shown as boxplots with median, interquartile range, and outliers. Numbers above each box indicate sample size. **b**, Diagnostic composition of key evaluation cohorts, including ADNI, NIFD, BraTS, ABIDE, and ADHD200, shown as proportions of controls and clinical groups. **c**, Relative contribution of each dataset to the full NeuroFM training corpus (n = 236,355 volumes), highlighting the dominance of large training (LDM100k) population (UKB) datasets alongside smaller disease-focused cohorts. **d**, Age distribution of the full dataset, grouped into decade bins. **e**, Sex distribution across all volumes.

#### Preprocessing

All datasets were processed through a uniform, minimal pipeline designed to preserve anatomical variability while ensuring cross-dataset compatibility. Raw MRI volumes were conformed to 1 mm isotropic resolution and centred within a 256^3^ voxel volume using FreeSurfer 7.4.1 [10]. Non-brain tissue was removed using SynthStrip [77]. During training and inference, voxel intensities were z-scored per volume using each image’s mean and standard deviation. No spatial registration or nonlinear normalization was applied. Processing in native space preserves genuine morphological variability and avoids the spatial and intensity distortions introduced by template-based registration, particularly in demographically diverse, paediatric, and pathological cohorts [78–82]. Any dataset-specific preprocessing steps are described explicitly where relevant.

### 4.2 Method 2: NeuroFM foundation model

NeuroFM is built using a novel 3D convolutional neural network architecture trained in a supervised manner (architecture details in Supplementary Fig. S2). The network is designed to be parameter-efficient, achieving representational capacity comparable to deep residual architectures while using fewer parameters. This is achieved through bottleneck compression blocks [9], which reduce the dimensionality of intermediate features without compromising depth or expressivity. Compared with conventional 3D ResNet-style architectures [33] of similar depth, this design enables improved computational efficiency and scalability for large-scale pretraining.

#### AI-generated (synthetic) supervision and training targets

Pretraining was performed exclusively on the LDM100k dataset [28, 29], a large-scale synthetic MRI resource generated by a generative AI model trained on UK Biobank data. The dataset comprises 100,000 anatomically realistic brain MRI volumes, evenly distributed across age and sex, and matched to the age range of the UK Biobank population. Using synthetic MRI data provides several advantages over traditional, real, brain data: an unlimited number of training samples can be generated, and there are minimal privacy, data governance, and anonymization concerns. Notably, LDM100k exhibits a uniform age distribution, avoiding the age-dependent sampling bias commonly observed in real cohorts.

#### Foundation model pretraining and model selection

NeuroFM was pretrained using multi-task supervised learning, with targets including brain age, biological sex, total brain volume, and ventricular volume. These complementary targets were selected to encourage the learning of global anatomical structure, demographic variability, and biologically meaningful morphological features within a single unified representation. The selected targets match the variables used in the synthetic MRI generation process. Training was conducted for 60 epochs (with early stopping) using an exponentially decaying learning rate. A large amount of data augmentation was applied to the input data, as the LDM100k data is quite uniform and free of many image artefacts. During pretraining, assorted augmentation transforms were applied to 95% of training volumes. The augmentations included three categories: geometric (e.g. translation and rotation), and intensity (e.g. noise and contrast changes), and artefacts (e.g. motion and ghosting) with varying levels of intensity and application probability depending on the specific augmentation (Supplementary Fig. S4; Supplementary Table ST5). Motion [83] and ghosting were applied from the TorchIO library [84], while inhomogeneity simulation was adopted from [13].

To assess the impact of model capacity on performance and generalizability, we trained three variants of NeuroFM with increasing parameter counts, denoted S (484,829 parameters), M (6,535,877 parameters), and L (10,815,109 parameters) models. The M-sized model achieved the best overall performance on the pretraining task (*R*^2^ = 0.86 patient age; *R*^2^ = 0.97 ventricular volume; Supplementary Table ST6). See Supplementary Table ST7 for the selected pretraining hyperparameters for each model size.

After pretraining, all three model variants were tested for finetuning sample efficiency and hyperparameter optimization over different training regimes as described in Method 5. Due to comparable finetuning performance across various training scenarios, the M model variant was selected for the UK Biobank analysis (Method 9) for compute efficiency. For the downstream task analyses (Method 6), the specific model variant used was selected based on superior performance characteristics of the downstream model. The selected model for each task is reported in the corresponding Method. Complete implementation details, code, and containers are available at https://github.com/rockNroll87q/NeuroFM.

### 4.3 Method 3: Predictor mode

NeuroFM was pretrained to predict brain age, biological sex, total brain volume, and ventricular volume (Method 2). In predictor mode, the pretrained network is used end-to-end, with all four outputs obtained directly by passing an MRI volume through the model. The four outputs obtained are used as features for downstream tasks. From the predicted age, the brain age gap (BAG) was computed as the difference between the predicted brain age and chronological age since it has been shown to serve as a sensitive biomarker of brain health, capturing individual deviations from normative ageing trajectories and relating to a wide range of clinical, social, and behavioural outcomes [18, 38, 39]. Unless otherwise stated, all downstream analyses involving age-related effects were performed using brain age gap (BAG) rather than raw predicted brain age.

### 4.4 Method 4: Encoder mode

Downstream analyses were performed using zero-shot feature extraction, where each MRI volume is passed through the pretrained network (Method 2), and the resulting latent representation is used directly for downstream analysis without any additional network training or fine-tuning. For each input volume, latent representations were extracted from the penultimate layer of NeuroFM, yielding fixed-length feature vectors of dimensionality 256 for the M model (161 for S; 512 for L). This resulted in a *N* × *D* feature matrix for each dataset, where *N* denotes the number of samples (i.e., volumes) and *D* the model-specific feature dimensionality.

These extracted features were used as inputs to machine-learning models to predict task-specific targets and evaluate performance against ground truth. All zero-shot analyses employed linear and logistic regressions for regression and classification tasks respectively (linear probing). Model evaluation was conducted using five-fold cross-validation, with folds grouped at the individual level to prevent data leakage across training and test sets. Statistical significance was assessed via permutation testing using 500 random label permutations. All t-SNE visualizations shown in Figs. 2, 3, and 5 were generated using features derived in encoder mode.

### 4.5 Method 5: Fine-tuning and retraining strategies

Although NeuroFM supports zero-shot inference (Methods 3 and 4), certain downstream tasks benefit from fine-tuning, defined as updating a subset of pretrained model parameters using task-specific supervision. In these cases, we retrained selected layers of the pretrained network described in Method 2. The task-specific output head was fully reinitialized with random weights and retrained to accommodate changes in the output target. Depending on the finetuning strategy, additional layers were unfrozen and updated from their pretrained values.

We evaluated two fine-tuning approaches: unfreezing selected pretrained layers, and reinitializing selected layers with random weights prior to training. Fine-tuning was assessed on the UK Biobank dataset across model sizes (S, M, L), training set sizes, and input image sequence (T1-weighted or T2-weighted MRI), to characterize scaling behaviour and data efficiency. Although NeuroFM was pretrained exclusively on T1-weighted MRI, fine-tuning on as few as 500–1,000 T2-weighted volumes yielded age-prediction performance approaching that of the pretrained model on T1-weighted data, indicating that the learned representations transfer to alternative structural contrasts (Supplementary Fig. S5). By contrast, training the NeuroFM architecture from scratch on UK Biobank T2-weighted data required at least 40,000 volumes for the larger model variants, and at least 5,000 volumes for the smallest variant, to achieve comparable performance.

Results from these analyses informed the selection of finetuning strategies and hyperparameters for subsequent experiments, including UK Biobank modelling (Method 9), the multi-dataset MRIQC task (Method 12), and the generation of retrained models for task relevance visualization (Method 8).

### 4.6 Method 6: Downstream tasks

Encoder-mode and competing methods’ features were corrected for acquisition-related confounds and age using double machine learning prior to classification or regression (Method 7). This was done to mitigate the influence of image artifacts and motion-related effects that are common in clinical neuroimaging data and that can provide shortcut cues for deep learning models [85]. Classification and regression analyses in this section were restricted to volumes with available (i.e., non-failed extraction) MRIQC features for confound correction and diagnostic labels, resulting in reduced sample sizes (Supplementary Table ST3). This correction regime was used for all tasks in Method 6. This procedure ensured that diagnostic discrimination reflected disease-related neuroanatomical variation rather than demographic or scanner-related artefacts.

a. **Clinical pathology diagnosis classification**. We assesed clinical classifications using the NIFD (Frontotemporal Lobar Degeneration Neuroimaging Initiative) and ADNI (Alzheimer’s Disease Neuroimaging Initiative) datasets. We used NIFD to evaluate the classification of frontotemporal dementia (FTD) versus healthy controls (HC), as well as FTD subtype differentiation. Using ADNI, we performed classification of Alzheimer’s disease (AD) versus HC, and multiclass classification of AD, mild cognitive impairment (MCI), and HC. For visualization, we applied t-SNE [86] to the encoder-model latent outputs (Method 3) to generate the low-dimensional embeddings shown in Fig. 2. All classification tasks were implemented using logistic regression (scikit-learn v1.3.2) [87] and evaluated under predictor mode (Method 3), encoder mode (Method 4), and competing methods (Method 13; Fig. 2). Attribution masks were generated using Method 8 to identify brain regions contributing to the classification decisions. LRP maps were generated for all four tasks in Fig. 2, but we present only the FTD subtype results in the main text. While the other task LRP maps do demonstrate some anatomical plausibility (Supplementary Fig. S16), it is possible that they were confounded by noise artefact differences between dementia-positive and healthy individuals.
b. **Glioma size prediction**. As an additional out-of-domain evaluation, we applied NeuroFM to glioma tumour size prediction and approximate tumour localization. Rather than performing tumour versus non-tumour classification, which can be confounded by site-specific acquisition artifacts and inter-dataset distributional differences (AUC=0.99; Supplementary Fig. S13), we focused on tumour size prediction as a more robust and biologically meaningful task. This formulation reduces the risk of trivial solutions driven by image quality or scanner-related cues and enables assessment of whether the model weights anatomically relevant brain regions. For visualization, we computed t-SNE projections using predictor-mode outputs, as shown in Fig. 2. Gliomas were dichotomized into small and large tumours based on a median split of tumour volume, and classification was performed using predictor mode (Method 3), encoder mode (Method 4), and competing methods (Method 13). Finally, attribution masks highlighting spatial regions contributing to tumour size predictions were generated using Method 8.
c. **Cognitive score prediction**. We performed cognitive analyses on ADNI and NIFD datasets using predictor mode outputs derived from NeuroFM (Method 3). Analyses focused on Clinical Dementia Rating (CDR) and Mini-Mental State Examination (MMSE) scores. Predicted brain age was obtained for each MRI volume and converted to brain age gap (BAG) by subtracting chronological age. In Fig. 3, column I, predicted versus chronological age is shown, with samples coloured by cognitive score to visualize the relationship between brain-age deviation and cognitive performance. Fig. 3, column II summarizes BAG distributions across increasing levels of cognitive impairment. For downstream inference, NeuroFM latent features were used to perform binary classification of cognitively unimpaired versus cognitively impaired individuals, with results reported in Fig. 3, columns III and IV. Classification was conducted using logistic regression (scikit-learn v1.3.2) [87] applied to the NeuroFM latent features.
d. **Neurodevelopmental disorder classification**. We conducted neurodevelopmental disorder analyses on the ADHD200 [52] and ABIDE [53, 54] datasets using features derived from NeuroFM. We used encoder-mode latent outputs to generate low-dimensional t-SNE projections for visualization (Fig. 5, column I). Diagnostic classification was performed using logistic regression on predictor mode (Method 3), encoder mode (Method 4), and competing methods (Method 13) features, with performance quantified as area under the receiver operating characteristic curve (AUC) (Fig. 5, column II) and corresponding confusion matrices reported in Fig. 5, column III. For ADHD200, we evaluated binary classification of ADHD versus controls, as well as subtype differentiation across control, hyperactive, combined, and inattentive groups using a one-vs-rest strategy. For ABIDE, we performed binary classification of autism spectrum disorder versus controls.

### 4.7 Method 7: Double machine learning for nuisance control

Acquisition- and quality-control-related artifacts can inadvertently encode diagnostic information, leading naïve models to achieve spuriously high performance in clinical classification tasks [68, 85]. In preliminary analyses, several baseline approaches, including a standard residual CNN [33], a competing deep architecture, and an MRIQC-only model, achieved unusually high apparent performance for frontotemporal dementia versus healthy control classification (AUC *>* 0.85; Supplementary Fig. S6-S7), suggesting strong confounding by acquisition characteristics rather than disease-related neuroanatomy. To address this, we applied a double machine learning (DML) procedure to control for artifact-related confounds, following established formulations [88, 89]. Importantly, DML was used as a conservative correction strategy, not as a performance-enhancing technique: it typically reduced classification performance while increasing the validity of model comparisons. This procedure was applied only to downstream discriminative analyses (e.g., classification tasks), and not to population-level regression or normative modelling.

DML estimates and subtracts feature components that can be explained by image quality metrics, producing artefact-controlled representations for subsequent analyses. Let *X* ∈ ℝ^*N×D*^ denote the z-scored feature matrix extracted from a model, with one row per MRI volume (*N*) and dimensionality *D* as 161, 256, or 512 dimensions, depending on model size. Let *Q* ∈ ℝ^*N×K*^ denote the matrix of quality-control variables derived from MRIQC, composed by CNR, SNR, and SNRd metrics. For each feature dimension *D* = 1, …, *D* dependence on quality-control variables was modelled as:

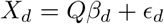

where *β*_*d*_ are artifact-related coefficients and *ϵ*_*d*_ are residuals. Coefficients *β*_*d*_ were estimated using ridge regression with L2 regularization:

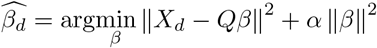

To prevent information leakage, nuisance parameters were estimated using fivefold cross-fitting, with folds grouped by individual and acquisition site. For each fold *f*, nuisance parameters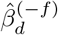were estimated on the remaining folds, and out-of-fold predictions were computed as:

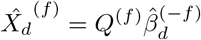

Artefact-controlled features were obtained as residuals:

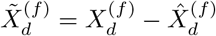

The resulting matrix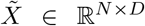constitutes the artefact-controlled feature representation used in subsequent analyses.

This approach produces approximately orthogonalized features that reduce sensitivity to linear quality-control-related variation. Given the high dimensionality of the feature space relative to the sample size, ridge regularization constrains the effective complexity of the nuisance model, and cross fitting reduces bias from reusing data.

Although artifact correction can remove variance correlated with true disease effects, this approach reduces over-optimistic bias driven by acquisition confounds and enables more meaningful model comparison. In the FTD classification task, baseline model performance dropped substantially after correction (AUC ≤ 0.7), whereas NeuroFM retained robust performance (AUC = 0.83; Fig. 2a), suggesting that its representations capture disease-relevant information beyond acquisition characteristics (Supplementary Fig. S6-S7).

### 4.8 Method 8: Interpretability and representation analyses

While comparative metrics and statistical analyses quantify task performance and generalization, they provide limited insight into which regions of the input drive model predictions. To obtain voxel-level relevance maps, we applied Layerwise Relevance Propagation (LRP) [30], an explainable method that redistributes prediction scores backward through the network to identify input features contributing to the output. We closely followed the implementation and configuration described by [4], with minimal modifications. LRP was computed on held-out test samples using models trained on the corresponding training splits. Post-processing of the resulting relevance volumes included co-registration to the MNI152 space, standardization (z-scoring), restriction to positive relevance values, and masking voxels outside the template brain tissue structures. For visualization, relevance maps were thresholded to the top 1–5% of values.

For all visualizations (Fig. 2c; Supplementary Fig. S16), we used parameters *α* = 2, *β* = 1, and *ϵ* = 0.25, emphasizing relevance contributions from the disease-positive class. Due to framework limitations, LRP was applied to functionally identical models instantiated without hierarchical layer abstractions, while preserving comparable connectivity and pretrained weights. All relevance maps were generated using finetuned NeuroFM-M models (Methods 2 and 5) trained for the corresponding target tasks, as LRP requires access to task-specific layer activations and gradients, which are not directly available in the zero-shot inference setting.

### 4.9 Method 9: Population-level socio-behavioural analyses

To examine socio-behavioural and clinical correlates of brain ageing, NeuroFM was fine-tuned on a healthy subset of the UK Biobank (n = 7,700) to predict brain age (Method 5), from which we derived brain age gap (BAG), the difference between predicted and chronological age. While the UK Biobank is generally restricted to healthy participants, there are some who have previous hospital-linked (ICD-10) or self-reported diagnoses. As such, our selected healthy subset selected only those free of diagnostic history related to our downstream targets (traumatic brain injury, neurocognitive illness, cardiovascular illness, etc.) and were additionally filtered to those with little to no smoking or drinking history.

Fine-tuning was performed to demonstrate adaptation of the foundation model to a new large-scale population dataset, rather than being used as a fixed feature extractor, in a comparison of two separate imaging modalities (T1w and T2w). For further comparison, we used NeuroFM-M was as a fixed BAG predictor without finetuning on the T1w images (Supplementary Fig. S15). Associations between BAG and clinical, cognitive, lifestyle, and demographic variables were evaluated in an independent held-out test set using fully adjusted linear regression models, independently for each experimental setting (T1w and T2w finetuned, T1w frozen). All models included age and sex as covariates, and predictors were standardized prior to analysis. Regression coefficients therefore represent standardized effect sizes, with positive values indicating factors associated with higher BAG (representing older-appearing brains relative to chronological age) and negative values indicating lower BAG (Fig. 4, col. I). To assess sex-specific effects, interaction models were fitted by including a sex and predictor interaction term for each variable. Interaction coefficients quantify differential associations between predictors and BAG in males versus females (Fig. 4, col. II). All models were estimated independently for each variable to reduce collinearity and improve interpretability. Statistical significance was assessed using two-sided tests, and false discovery rate (FDR) correction was applied across all tested variables using the Benjamini–Hochberg procedure (q *<* 0.05). Effect estimates and associations were computed on the held-out test set only, ensuring that association analyses were not influenced by fine-tuning data.

### 4.10 Method 10: Normative modelling

We constructed normative models using predictor outputs from NeuroFM (Method 3) derived from cognitively normal T1-weighted MRI volumes from the ADNI dataset. Models were trained exclusively on cognitively normal individuals and subsequently applied to study individuals to quantify deviations from normative ageing trajectories. Sex-stratified normative trajectories were estimated for BAG, total brain volume, and ventricular volume to account for known sex-related differences in brain morphology. We performed normative modelling using a second-order polynomial regression model implemented in PCNToolkit [25] without explicit modelling of within-individual dependence, yielding estimates of the expected mean and age-dependent variance. A second-order form was selected to capture non-linear age effects while avoiding over-fitting (normative curves shown in Supplementary Fig. S9). Individuals analysed in Fig. 7 were held out and not used to fit the normative models. Individual observations were mapped onto the normative reference distribution to derive standardized deviation scores (z-scores) relative to age-matched controls. Models were restricted to the age range 50–95 years. Direct comparisons between NeuroFM and FreeSurfer predictions are reported in Supplementary Fig. S10. Results are reported in Fig. 7 and Extended Data Fig. 2.

### 4.11 Method 11: Longitudinal disease risk estimation

We estimated longitudinal disease risk using sex-stratified Cox proportional hazards models [56] from the lifelines package (https://lifelines.readthedocs.io/), constructed on an age-corrected basis using outputs from the normative models (Method 10). Survival time was defined as the interval from baseline assessment to clinical conversion, where survival corresponded to remaining free of the target diagnosis during follow-up (e.g., survival meaning not diagnosed with Alzheimer’s dementia). Three independent time-to-event models were fitted to capture distinct disease transitions: healthy to mild cognitive impairment (MCI), healthy to Alzheimer’s disease (AD), and MCI to AD. For each model, covariates included normative z-scores derived from NeuroFM for BAG, total brain volume, and ventricular volume, together with chronological age and sex at baseline. The partial effects models show a wide distribution of survival outcomes depending on covariate and the model used (Supplementary Fig. S11), with a maximum outcome timeline of 14 years.

Cox proportionality assumptions held for all covariates, as verified by Supplementary Fig. S12. The covariate hazard ratios for the healthy to AD model were BAG=0.45 ([0.32, 0.58] 95% CI), ventricle volume=0.17 ([0.05, 0.28] 95% CI), age=0.04 ([0.02, 0.06] 95% CI), and total brain volume=-0.63 ([-0.77, -0.49] 95% CI). For the MCI to AD model, they were BAG=0.36 ([0.23, 0.49] 95% CI), ventricle volume=0.16 ([0.04, 0.28] 95% CI), age=0.06 ([0.04, 0.08] 95% CI), and total brain volume=-0.60 ([-0.75, -0.45] 95% CI).

As with the normative models, individuals included for the analysis in Fig. 7 and Extended Data Fig. 2 were held out from the training data for the fitted Cox models. We computed individual-level risk estimates from the fitted survival functions, with 5-year risk calculated based on baseline covariates at each examination. For longitudinal case studies, risk trajectories were updated at each follow-up visit using time-varying baseline information. The update at each step was performed using the most relevant time-to-event model based on the most recent diagnosis at or before that visit (e.g. an individual with MCI status would have their risk estimated using the MCI to AD fitted Cox model). Individuals were grouped into four subtypes: those who were cognitively normal throughout the study period (stable HC), those who were MCI throughout the study period (stable MCI), those who progressed from cognitively normal to MCI, and those who progressed from cognitively normal to AD.

### 4.12 Method 12: Quality metric prediction and artefact detection

#### a Quality metric prediction from MRIQC

To build a deep learning model that emulates MRIQC [55] and disentangles quality-related artefacts from the latent representations, we adapted NeuroFM (Method 2) using both zero-shot regression (Method 4) and finetuning (Method 5). These approaches were evaluated empirically to determine which yielded superior prediction of MRIQC-derived quality metrics.

We computed ground-truth MRIQC values using the MRIQC pipeline (https://github.com/nipreps/mriqc). Although MRIQC was applied to all available volumes, several datasets exhibited high failure rates, which limited subsequent analyses (Supplementary Table ST1). Successful outputs were used as training targets for this method and as inputs for debiased machine learning (Method 7) in downstream analyses (Method 6).

Models were trained to predict the following subset of MRIQC metrics, selected to span signal-to-noise, contrast, spatial smoothness, intensity non-uniformity, tissue composition, and artefact-related measures: cjv, cnr, snr csf, snr gm, snr total, snrd csf, snrd gm, snrd wm, snr wm, efc, fber, inu med, inu range, fwhm avg, icvs csf, icvs gm, icvs wm, qi 2, summary wm mean, summary gm mean, summary csf mean, and summary bg mean (definitions in Supplementary Table ST2). QI1 was excluded due to a high rate of invalid or missing values across datasets. Datasets and training/testing splits are shown in Supplementary ST4.

#### b Motion artefact detection

We assessed motion artefact detection using the Autism Brain Imaging Data Exchange dataset (ABIDE; n = 1,010) [53, 54]. Synthetic artefacts were introduced to emulate ghosting and head motion. Augmentations were applied per volume using TorchIO image transforms [84]. Each volume was augmented with motion (35% probability), ghosting (35%), and one additional randomly sampled transform from the augmentation library (100%). Some volumes had more than one target augmentation applied simultaneously, and this was used for the joint classification task.

For classifier training, latent features were extracted from NeuroFM and comparative models (Method 13). Logistic regression models were trained to predict artefact labels from these features using fivefold cross-validation with held-out folds for evaluation (Method 4). Dataset partitions were grouped by individual to prevent leakage between training and testing sets.

In parallel, MRIQC was applied to the augmented volumes, and logistic regression models were trained on the resulting quality metrics to predict artefact labels. The 3D convolutional neural network trained from scratch (Method 13.a) was first pretrained on an independent set of augmented volumes generated with the same augmentation probabilities, then used as a feature encoder for downstream classification.

We quantified model performance using the AUC under three evaluation conditions: ghosting only, motion only, and combined ghosting plus motion. All reported NeuroFM results from Method 12.a-b use NeuroFM-S (Method 2; Fig. 6). The M and L variants either failed to converge or showed reduced performance in quality control prediction (Supplementary Fig. S1). Additional analyses are reported in the Supplementary Materials.

### 4.13 Method 13: Comparative methods

a. **ResNet-style convolutional neural network**. As a naïve deep learning baseline, we used a 3D convolutional neural network (3D CNN) with the same layer architecture and parameter count as NeuroFM-M (6,535,877 parameters). We refer to this as ResNet-style [33] since NeuroFM is based on a residual network architecture. When used as a comparator (Method 6) the 3D CNN was initialized with random weights and fully trained on the same dataset samples, using identical data splits, cross-validation folds, and output targets. For example, the 3D CNN was trained to predict AD vs healthy cognitive status in Fig. 2a.
b. **Simple Fully Convolutional Network (SFCN)**. We used the SFCN architecture described in [19] and [90], a lightweight 3D CNN originally developed for brain age prediction and subsequently applied to a range of neuroimaging tasks, including dementia prediction [4]. To enable a controlled architectural comparison, we pretrained the SFCN architecture from scratch using the same pretraining data and protocol as NeuroFM-M, selecting the model variant with a comparable parameter count. As SFCN was designed as a brain age prediction model, the age of each individual was set as the output training target during pretraining. The SFCN architecture was originally designed with an input shape of 160 × 192 × 160 voxels, to accommodate the MNI-registered input volumes. To ensure a full comparison with our own architecture, this was updated to 256^3^ to match NeuroFM. For downstream evaluations, SFCN was applied using the same inference and finetuning strategies as NeuroFM. Specifically, when NeuroFM was evaluated in a linear inference setting (Methods 3 and 4), the same protocol was applied to SFCN. When finetuning was performed (Methods 5 and 6), we matched training procedures, hyperparameters, and layer adaptation strategies as closely as possible. Although NeuroFM’s latent dimensionality varies from 161 to 512 features depending on model size (see Method 2), SFCN uses a fixed 64-dimensional latent representation.
c. **Brain Imaging Adaptive Core (BrainIAC)**. Brain Imaging Adaptive Core (BrainIAC) [22] is a self-supervised brain MRI foundation model designed to learn generalizable representations from unlabelled data for adaptation across diverse downstream tasks. The model uses a Vision Transformer Base (ViT-B) encoder and was pretrained using contrastive self-supervised learning on template-registered brain MRIs drawn from datasets spanning ten neurological conditions. Unlike SFCN, which was designed around a single pretraining target (brain age), BrainIAC was pretrained without any specific output label, instead learning representations through positive and negative pair contrast. For downstream evaluations in this study, BrainIAC was applied using their provided inference pipeline to produce extracted latent features, which were then evaluated using linear probing (Methods 4 and 6). BrainIAC uses a fixed 768-dimensional latent representation, which is comparable to the NeuroFM-L variant.

### 4.14 Method 14: Longitudinal brain age trajectory analysis in stable and progressive MCI

We evaluated Longitudinal cortical trajectories preceding conversion to Alzheimer’s disease by adapting elements of a previously described framework [4]. Visit timelines were anchored to each individual’s conversion date for progressive MCI cases, or to the final available visit for stable individuals. To improve comparability of follow-up patterns, individuals were paired using the Hungarian algorithm, which minimized symmetric nearest-neighbour distances between negative-time visit schedules with a maximum allowable separation of 365 days.

Deviations from healthy ageing were quantified for each target feature using age-and sex-matched reference values derived from cognitively healthy controls. For each MCI individual, the nearest age match within sex was identified, and deviation scores were calculated as the difference between the observed value and the corresponding normative mean. These deviation measures were subsequently used as predictive features, with primary analyses focusing on the brain age gap.

We estimated feature trajectories using linear mixed-effects models with participant-specific random intercepts and slopes (measure ∼ time × group + age + sex). Time was centred at the clinical event, defined as either conversion or last recorded visit (0 years). Fixed effects captured baseline group differences, longitudinal change, overall group effects, and group-by-time interactions.

For visualization, visits were aggregated into one-year intervals with a minimum of 20 observations per bin. Group-level means and 95% confidence intervals were computed for each interval, and trajectories were displayed using bubble plots in which marker size reflected the number of observations contributing to each estimate. Full fitted model coefficients were *β*_0_ = 24 (*p <* 1 × 10^*−*10^), *β*_*time*_ = 0.398 (*p <* 1 × 10^*−*5^), *β*_*group*_ = 1.844 (*p <* 2 × 10^*−*3^), *β*_*time×group*_ = 0.13 (*p* = 0.26). Results are presented in Extended Data Fig. 2a.

## Data Availability

All data used in the manuscript are openly available online.

https://rocknroll87q.github.io/NeuroFM/

## Acknowledgements

We acknowledge the MVLS Advanced Research System (MARS) at the University of Glasgow for providing high-performance computing resources and technical support.

Some data used in the preparation of this article were obtained from the Alzheimer’s Disease Neuroimaging Initiative (ADNI) database. As such, the investigators within the ADNI contributed to the design and implementation of ADNI and/or provided data but did not participate in analysis or writing of this report. A complete listing of ADNI investigators can be found at: https://adni.loni.usc.edu/wpcontent/uploads/how_to_apply/ADNI_Acknowledgement_List.pdf. For up-to-date information, see adni.loni.usc.edu.

Some of the data used in the preparation of this article were obtained from the Neuroimaging in Frontotemporal Dementia (NIFD) dataset, part of the Frontotemporal Lobar Degeneration Neuroimaging Initiative (FTLDNI). Data collection and sharing for this project was funded by the Frontotemporal Lobar Degeneration Neuroimaging Initiative (National Institutes of Health Grant R01 AG032306). The study is coordinated through the University of California, San Francisco, Memory and Aging Center.

FTLDNI data are disseminated by the Laboratory for Neuro Imaging at the University of Southern California. For up-to-date information on participation and protocol, see http://memory.ucsf.edu/research/studies/nifd.

Data were provided [in part] by the Human Connectome Project, WU-Minn Consortium (Principal Investigators: David Van Essen and Kamil Ugurbil; 1U54MH091657) funded by the 16 NIH Institutes and Centers that support the NIH Blueprint for Neuroscience Research; and by the McDonnell Center for Systems Neuroscience at Washington University.

Data were provided [in part] by OASIS-3: Longitudinal Multimodal Neuroimaging: (Principal Investigators: T. Benzinger, D. Marcus, J. Morris); NIH P30 AG066444, P50 AG00561, P30 NS09857781, P01 AG026276, P01 AG003991, R01 AG043434, UL1 TR000448, R01 EB009352. AV-45 doses were provided by Avid Radiopharmaceuticals, a wholly owned subsidiary of Eli Lilly.

Data were [in part] obtained from the IXI dataset (https://braindevelopment.org/ixi-dataset/).

Data were [in part] provided by the 1000 Functional Connectomes Project (FCP).

For access and usage information, see https://fcon_1000.projects.nitrc.org.

## Declarations

### Funding

A. Dibble was supported by a PhD grant from the Scottish Graduate School of Social Science, Doctoral Training Partnership (SGSSS-DTP), on behalf of the Economic and Social Research Council (ESRC, grant number: ES/P000681/1).

C. Dalby was supported by a PhD grant by the Medical Research Council (MRC) as part of the Precision Medicine Doctoral Training Programme.

A.Fracasso was supported by a grant from the Biotechnology and Biological Sciences Research Council (BBSRC, grant number: BB/S006605/1) and the Bial Foundation (Bial Foundation Grants Programme; Grant id: A-29315, number: 203/2020, grant edition: G-15516).

This research has been conducted using the UK Biobank Resource under Application 17689.

### Author contributions (CRediT)

**Austin Dibble**: Conceptualization, Methodology, Software, Validation, Formal Analysis, Investigation, Data Curation, Writing – Original Draft, Writing – Review & Editing, Visualization. **Connor Dalby**: Conceptualization, Methodology, Validation, Formal Analysis, Writing – Review & Editing, Data Curation, Visualization. **Michele Sevegnani**: Conceptualization, Writing – Review & Editing, Supervision, Funding Acquisition. **Alessio Fracasso**: Conceptualization, Methodology, Writing – Review & Editing, Supervision. **Donald M. Lyall**: Conceptualization, Formal Analysis, Validation, Methodology, Resources, Writing – Review & Editing, Data Curation. **Monika Harvey**: Conceptualization, Writing – Review & Editing, Writing - Original Draft, Supervision, Funding Acquisition. **Michele Svanera**: Conceptualization, Investigation, Methodology, Software, Writing - Original Draft, Writing – Review & Editing, Data Curation, Visualization, Resources, Supervision, Funding Acquisition, Project Administration.

### Code availability

https://github.com/rockNroll87q/NeuroFM

## 5 Extended Data

**Extended Data Fig. 1.**
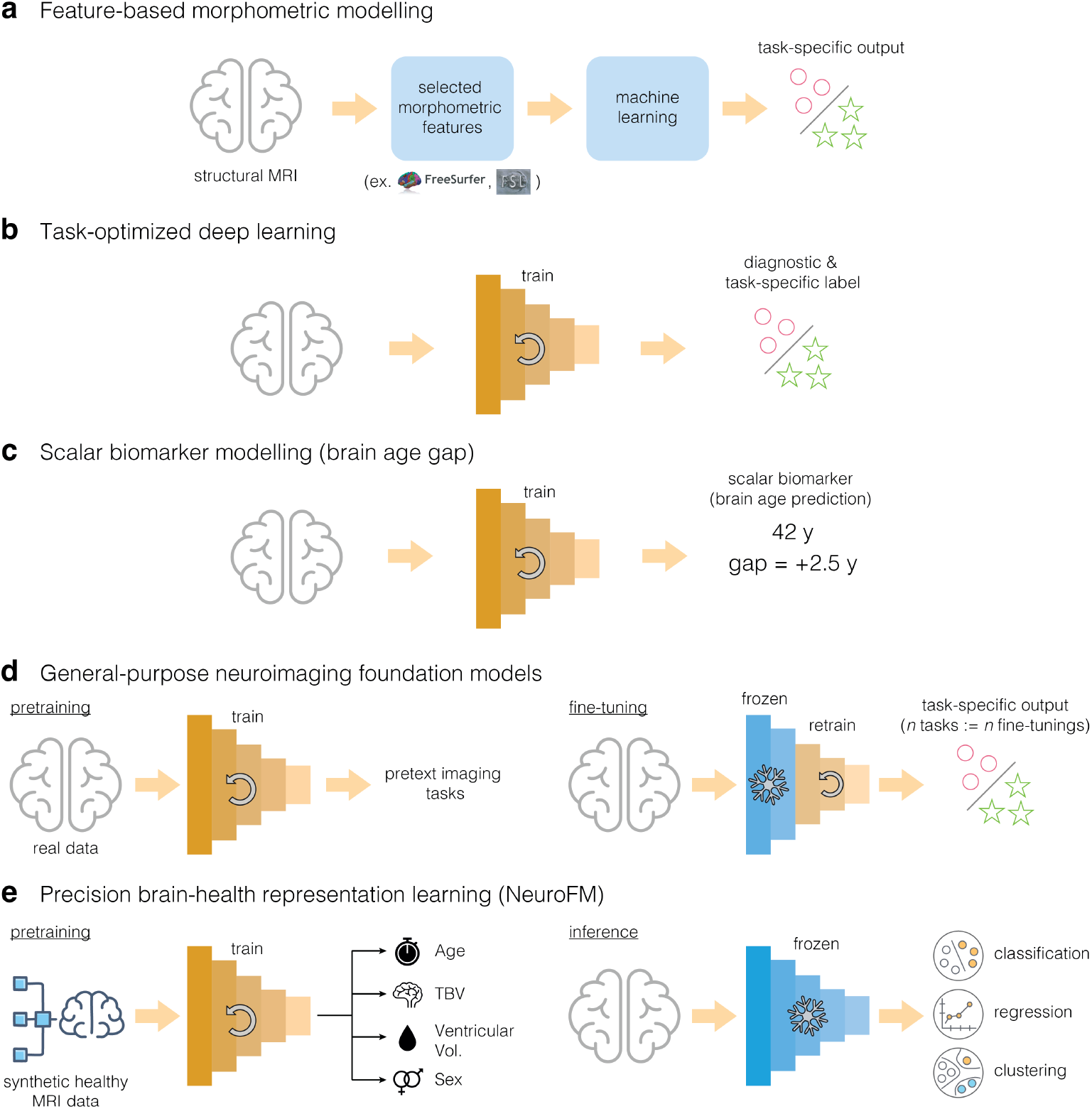
Evolution of structural MRI modelling paradigms toward precision neuroimaging. **a**, Feature-based morphometric modelling using hand-crafted anatomical features and classical machine learning. **b**, Task-optimized deep learning, which improves performance by learning task-specific image features but limits inference to single labels or cohorts. **c**, Scalar biomarker modelling (e.g., brain age gap), introducing normative reference but collapsing brain health into a single measure. **d**, General-purpose neuroimaging foundation models, where large-scale pretraining improves robustness but typically requires task-specific fine-tuning. **e**, Precision brain-health representation learning (NeuroFM), which learns a frozen, multidimensional representation of brain health from synthetic MRI and enables direct, person-level inference across domains using simple readouts.

**Extended Data Fig. 2.**
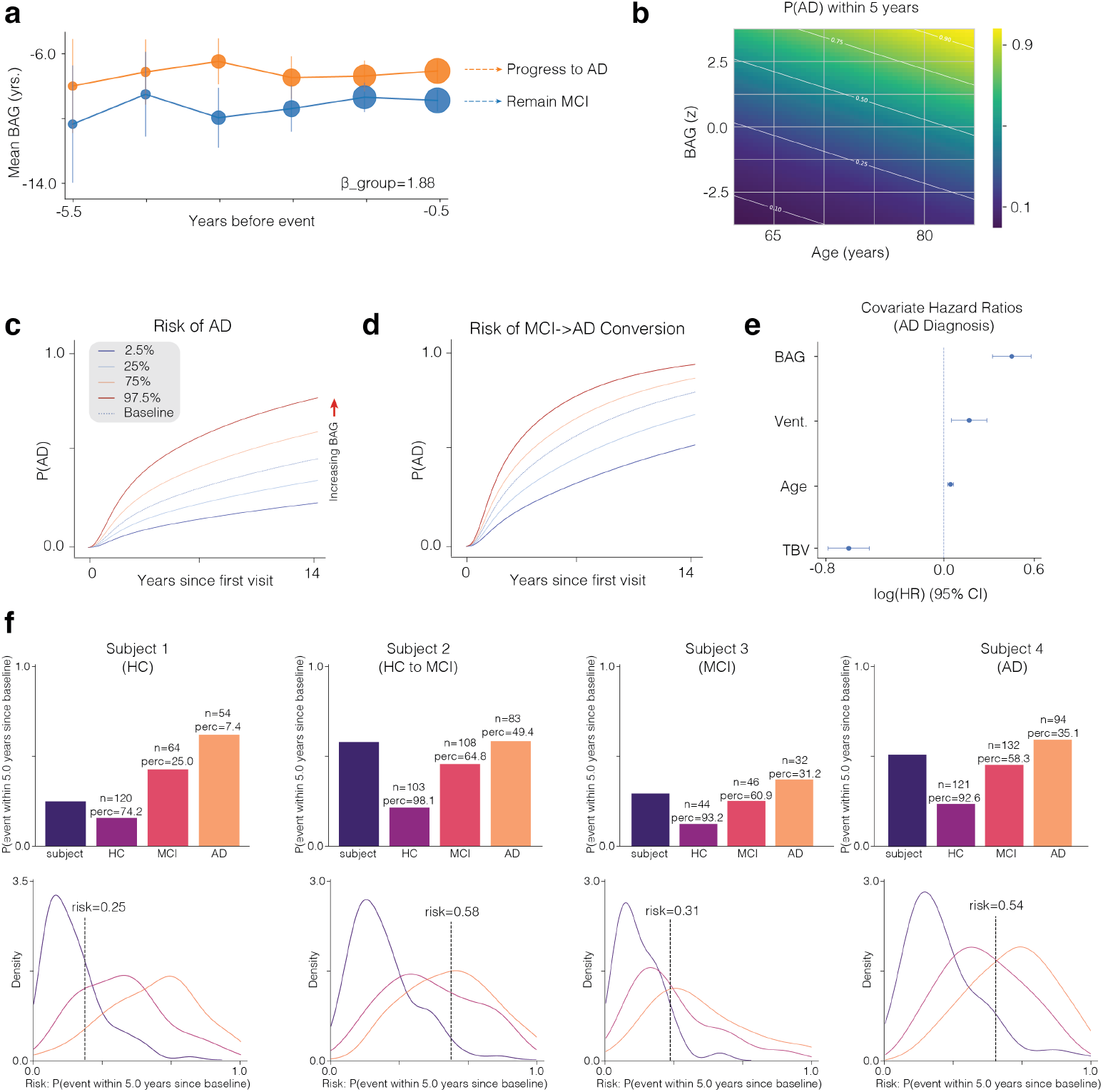
Survival model projections underlying precision neuroimaging–based risk prediction. . **a**, Mean brain age gap (BAG) in individuals who progressed from MCI to AD versus non-progressors across a 5-year lookback period from the final diagnostic event or visit. Linear mixed-effects models (Method 14; group *β* = 1.88; *p* = 0.0019), indicate elevated BAG up to 5.5 years before conversion. **b**, Two-dimensional projection of the Cox proportional hazards model [56] estimating a 5-year risk of conversion from cognitively normal status to AD as a function of age and normative BAG z score; covariates fixed at male sex, brain volume -1 z, and ventricular volume 0 z. Risk increases with both variables. **c**, Predicted cumulative probability of AD conversion from a cognitively normal status to AD based on partial BAG effects; curves span the 2.5th to the 97.5th percentiles and show increasing probability with higher BAG. Values are group partial effects predicted by the fitted Cox models. **d**, Predicted cumulative probability conversion from MCI to AD derived from the same Cox framework as in panel c, again increasing with BAG. **e**, Covariate hazard ratios for conversion from cognitively normal status to AD, shown on a log scale with 95% confidence intervals. **f**, Individual 5-year probabilities of AD diagnosis compared with age- and sex-matched individuals across diagnostic groups (top); kernel density estimates illustrate each individual’s position within the matched probability distributions (bottom). Individuals correspond to Fig. 7

## Notes

### Competing Interest Statement

The authors have declared no competing interest.

### Author Declarations

Some data used in the preparation of this article were obtained from the Alzheimer's Disease Neuroimaging Initiative (ADNI) database. As such, the investigators within the ADNI contributed to the design and implementation of ADNIand/or provided data but did not participate in analysis or writing of this report. A complete listing of ADNI investigators can be found at: https://adni.loni.usc.edu/wp-content/uploads/how_to_apply/ADNI_Acknowledgement_List.pdf. For up-to-date information, see adni.loni.usc.edu. Some of the data used in the preparation of this article were obtained from the Neuroimaging in Frontotemporal Dementia (NIFD) dataset, part of the Frontotemporal Lobar Degeneration Neuroimaging Initiative (FTLDNI). Data collection and sharing for this project was funded by the Frontotemporal Lobar Degeneration Neuroimaging Initiative (National Institutes of Health Grant R01 AG032306). The study is coordinated through the University of California, San Francisco, Memory and Aging Center. FTLDNI data are disseminated by the Laboratory for Neuro Imaging at the University of Southern California. For up-to-date information on participation and protocol, see http://memory.ucsf.edu/research/studies/nifd. Data were provided [in part] by the Human Connectome Project, WU-Minn Consortium (Principal Investigators: David Van Essen and Kamil Ugurbil; 1U54MH091657) funded by the 16 NIH Institutes and Centers that support the NIH Blueprint for Neuroscience Research; and by the McDonnell Center for Systems Neuroscience at Washington University. Data were provided [in part] by OASIS-3: Longitudinal Multimodal Neuroimaging: (Principal Investigators: T. Benzinger, D. Marcus, J. Morris); NIH P30 AG066444, P50 AG00561, P30 NS09857781, P01 AG026276, P01 AG003991, R01 AG043434, UL1 TR000448, R01 EB009352. AV-45 doses were provided by Avid Radiopharmaceuticals, a wholly owned subsidiary of Eli Lilly. Data were [in part] obtained from the IXI dataset (https://brain-development.org/ixi-dataset/). Data were [in part] provided by the 1000 Functional Connectomes Project (FCP). For access and usage information, see https://fcon_1000.projects.nitrc.org. All datasets were de-identified by the data providers prior to our access. We did not have access to any personally identifiable information, such as names or addresses, at any stage of the study. Participants are represented only by anonymised subject identifiers assigned by the dataset curators (ex. subject-0021), and the linkage between identifiers and personal information is maintained solely by the original data custodians.

### Summary of Updates

Update the manuscript text after a round of internal revision.

