## Supplementary material for "NeuroFM: Toward Precision Neuroimaging with Foundation Models for Individualized Brain Health Estimation": supp

#### 1 Supplementary Figures

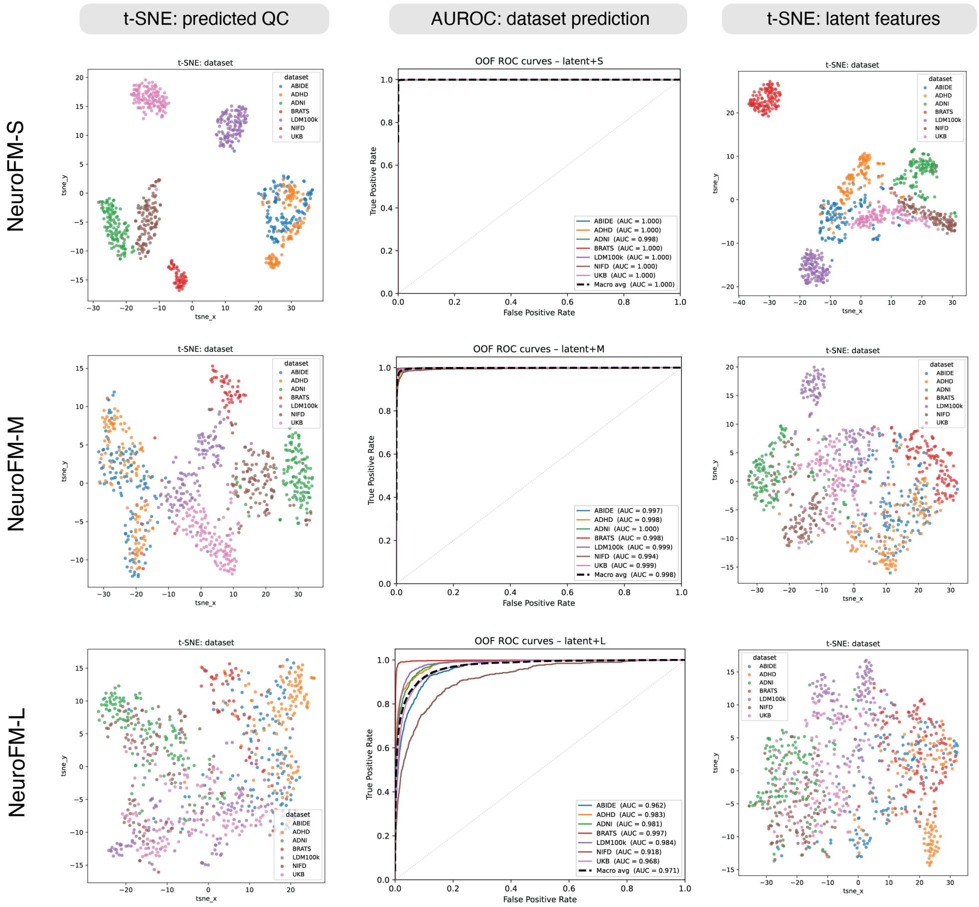


**Supplementary Fig. S1 | QC feature predictability as a function of NeuroFM model capacity.** Linear regression was used to predict MRIQC image quality features from NeuroFM representational features, in order to assess the degree to which quality-related variance was linearly disentangled from the learned representations. This analysis was performed across all three NeuroFM capacity variants (S, M, and L). **Left column:** t-SNE visualizations of predicted MRIQC features for each capacity variant, coloured by dataset cohort. Distinct cohort clustering observed in the S variant became progressively more diffuse in the M and L variants, indicating reduced linear separability of quality-related information with increasing model capacity. **Middle column:** logistic regression models were trained on NeuroFM features to predict dataset cohort directly as a classification task. Cohort identity was strongly recoverable across all capacity variants, though macro-average AUC declined modestly from 0.99 in the S and M variants to 0.97 in the L variant. **Right column:** t-SNE applied directly to raw NeuroFM extracted features revealed similar cohort-level clustering, which also became more diffuse in the M and L variants. Together, these results suggested that larger NeuroFM variants progressively abstracted away cohort-specific and quality-related signal, consistent with improved representational disentanglement at higher model capacity.

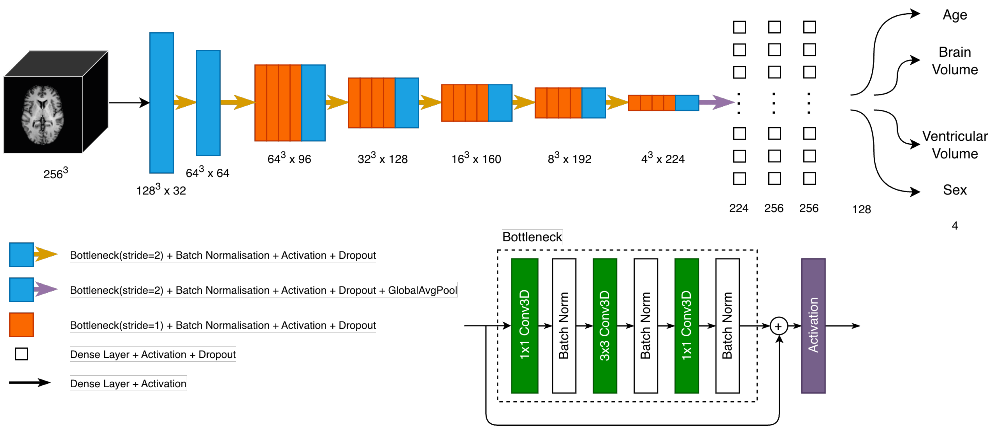


**Supplementary Fig. S2 | NeuroFM architecture diagram.** Basic architectural diagram for NeuroFM's M-size encoder CNN. A full $256^3$ MRI volume is input and passed through a series of "Bottleneck" CNN blocks, making up the model encoder. After the encoding step, global average pooling is used to map the activations to a series of fully connected layers. The number of fully connected layers used, and their sizes, depends on the network variant. Each Bottleneck layer is constructed of $1 \times 1 \times 1$ input and output convolutions, with a $3 \times 3 \times 3$ compression layer in the middle to reduce parameter sizes and force the model to derive a compressed representation of the input data. Batch normalization occurs after each convolution layer. A residual connection is passed from the input to the output before final activation.

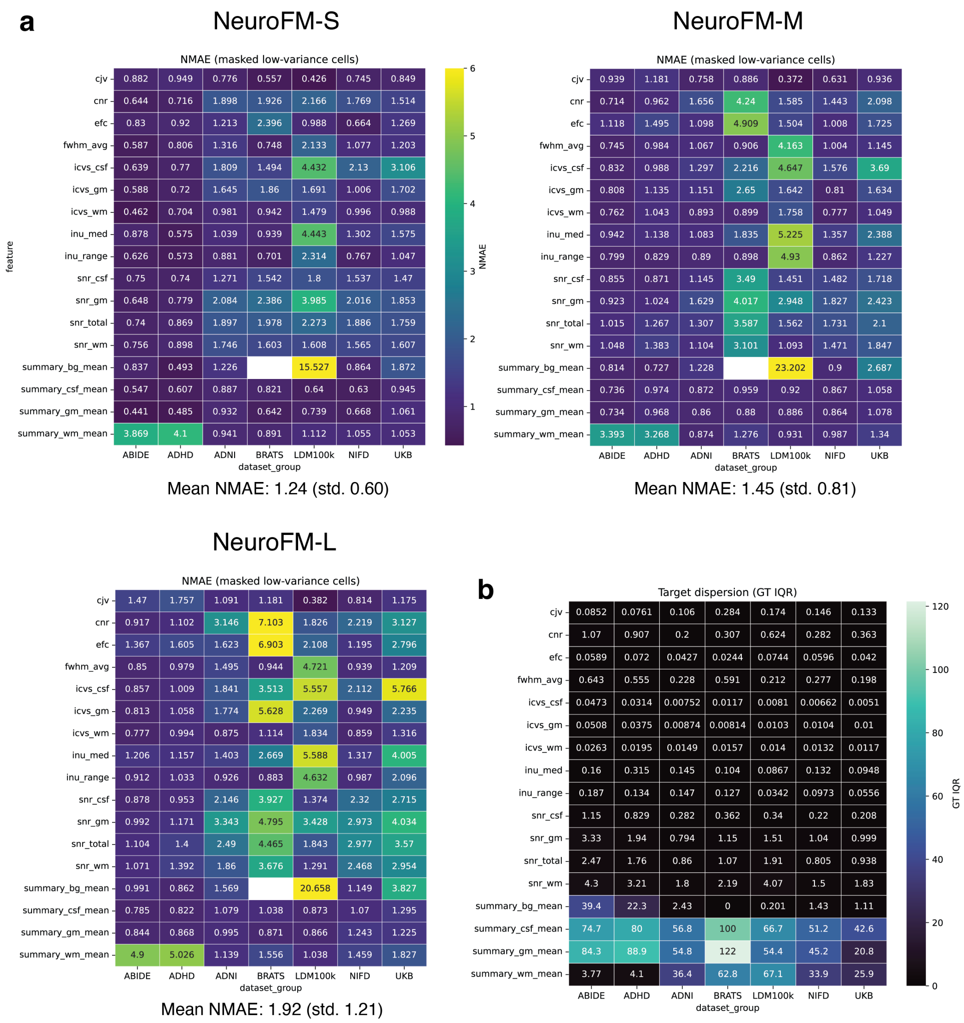


**Supplementary Fig. S3 | Prediction of MRIQC image quality features from NeuroFM representations across model capacity variants.** Linear regression models were trained on NeuroFM representational features to predict MRIQC-derived image quality metrics, and prediction accuracy was assessed across all three model capacity variants (S, M, and L). **a,** Normalized mean absolute error (NMAE) was calculated for each predicted quality metric within each dataset cohort, and results are displayed as a heatmap per NeuroFM variant. The summary_bg_mean feature was excluded from analysis in BraTS-2023 due to zero variance in its ground-truth distribution. Mean NMAE and standard deviation are reported for each variant. Mean NMAE increased with model capacity, indicating that larger variants were less linearly predictable from quality-related features, consistent with progressive disentanglement of artefact-related signal from the learned representations. **b,** Standardized interquartile range (IQR) is shown for each quality metric and dataset cohort combination, providing a measure of the distributional spread of predicted values relative to ground truth.

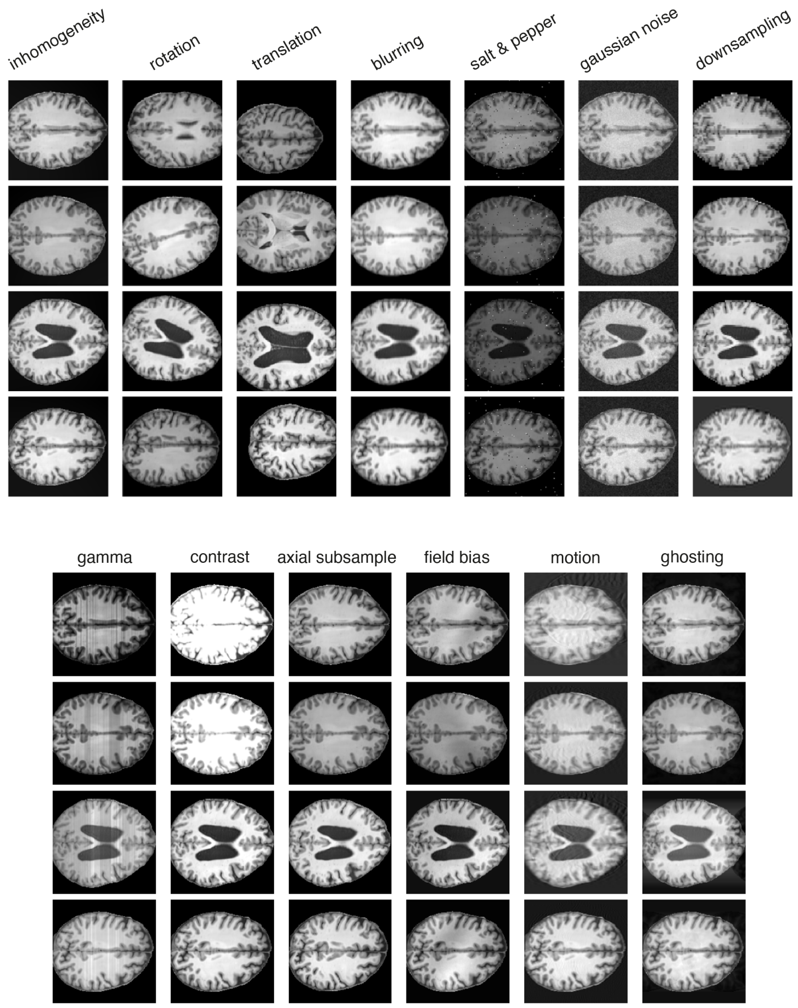


**Supplementary Fig. S4 | Augmentation examples.** Examples from our augmentation pipeline. All images are from the same volume in $256^3$ voxel space, sliced to give an axial view from the middle of the volume.

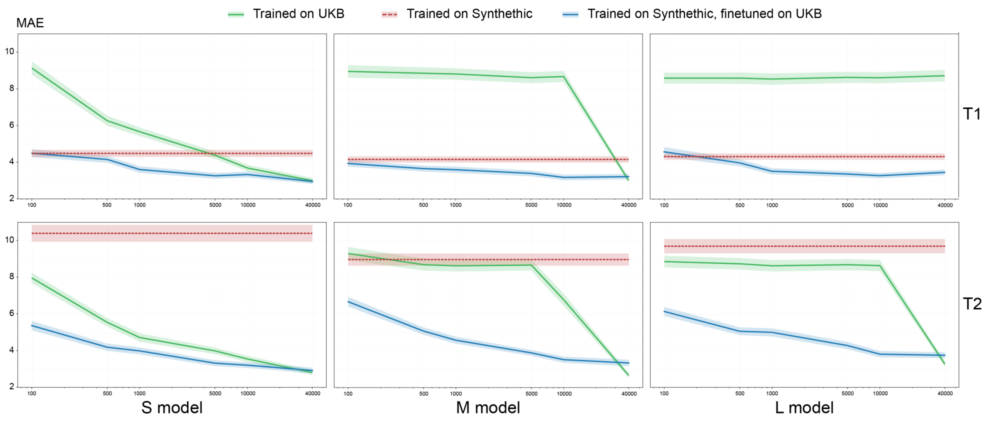


**Supplementary Fig. S5 | NeuroFM fine-tuning performance across MRI sequence, dataset size, and model capacity.** The utility of NeuroFM's pretrained representations was benchmarked against training equivalent models from scratch on the UK Biobank dataset, using brain age prediction (chronological age from scan) as the evaluation task. Both training regimes were evaluated across a range of dataset sizes (100 to 40,000 scans, the full single-sequence capacity of UK Biobank), two MRI sequences (T1-weighted and T2-weighted), and all three NeuroFM model capacity variants (S, M, and L). Performance was reported as mean absolute error (MAE) in years. In all panels, the **green line** denotes validation MAE for models trained from scratch; the **red line** denotes validation MAE of the pretrained model evaluated on a held-out validation set of 250 scans; and the **blue line** denotes validation MAE for fine-tuned NeuroFM models. For T1-weighted training, scratch-trained models required at least 5,000 scans to match pretrained validation MAE, and at least 10,000 scans to match fine-tuned MAE. For T2-weighted training, a sequence not seen during NeuroFM pretraining, scratch-trained models did not reach fine-tuned MAE until the full 40,000-scan dataset was used. This cross-sequence generalization demonstrated that NeuroFM's pretrained representations transferred efficiently to unseen MRI contrasts, substantially reducing the data required to achieve competitive performance relative to training from scratch.

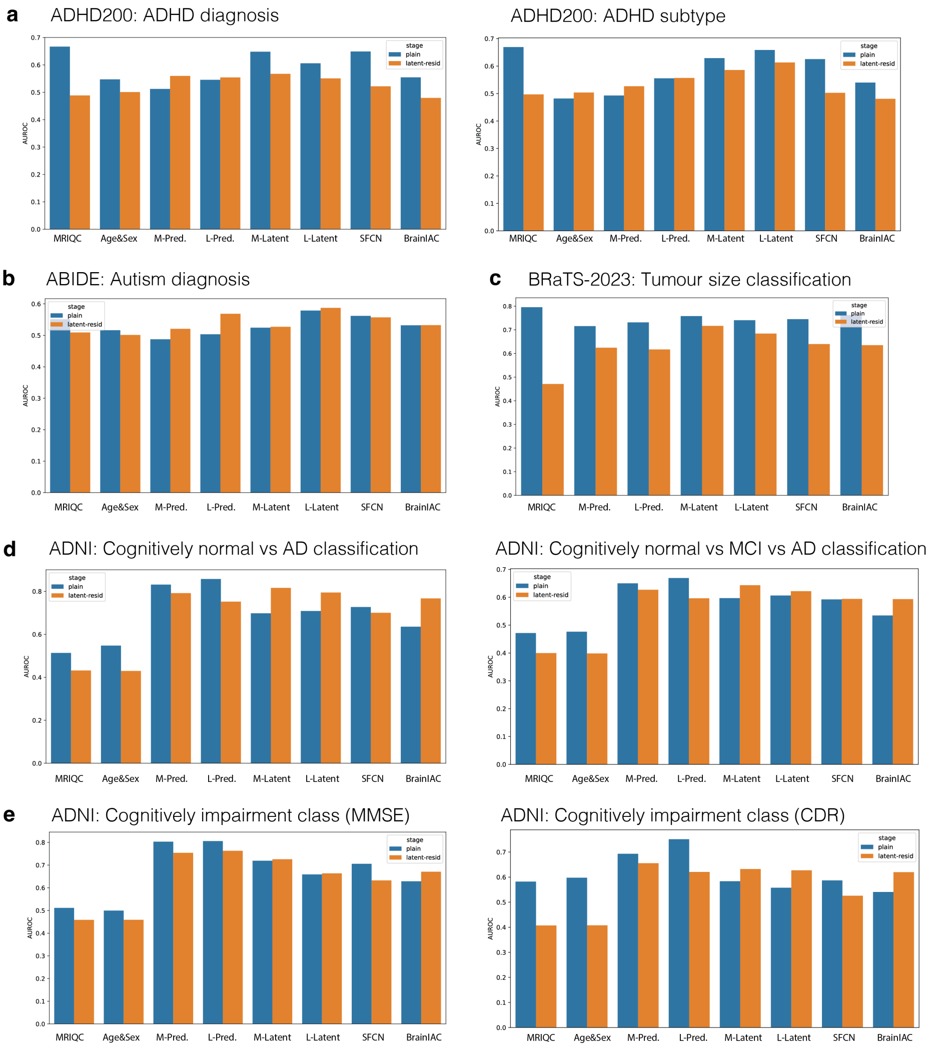


**Supplementary Fig. S6 | Effect of QC residualization on task AUROC across models.** Bar plots show AUROC of logistic regression classifiers trained on features extracted before (blue) and after (orange) quality-control (QC) residualization, applied to mitigate the influence of acquisition and demographic confounds on learned representations. Each group of bars along the x-axis corresponds to a distinct feature set used as classifier input. NeuroFM variants are denoted as follows: M and L indicate model scale (medium and large, respectively); Pred denotes features derived from the NeuroFM predictor head; Latent denotes features extracted from the NeuroFM encoder. SFCN and BrainIAC are included as external baseline models. **a,** ADHD-200 cohort: ADHD binary classification (left) and ADHD subtype classification (right). **b,** ABIDE cohort: autism spectrum disorder (ASD) binary diagnosis classification. **c,** BraTS-2023 cohort: binary classification of tumour size. **d,** ADNI cohort: cognitively normal (CN) versus Alzheimer's disease (AD) binary classification (left), and CN versus mild cognitive impairment (MCI) versus AD three-class classification (right). **e,** ADNI cohort: binary cognitive impairment classification using Mini-Mental State Examination (MMSE; left) and Clinical Dementia Rating (CDR; right) scores.

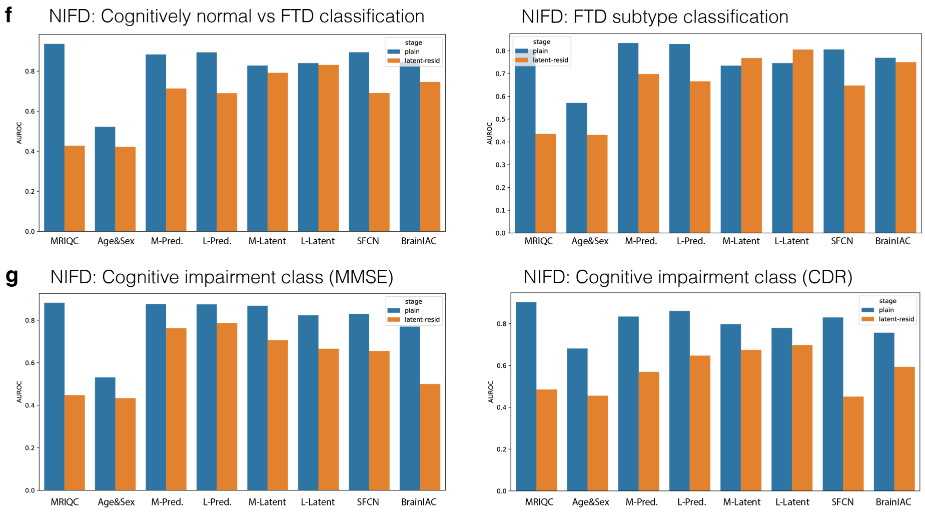


**Supplementary Fig. S7 | Effect of QC residualization on task AUROC across models.** Continuation of Supplementary Fig. S6. All display conventions are as described therein. **f,** NIFD cohort: CN versus frontotemporal dementia (FTD) binary classification (left), and FTD subtype classification (right). **g,** NIFD cohort: binary cognitive impairment classification using MMSE (left) and CDR (right) scores.

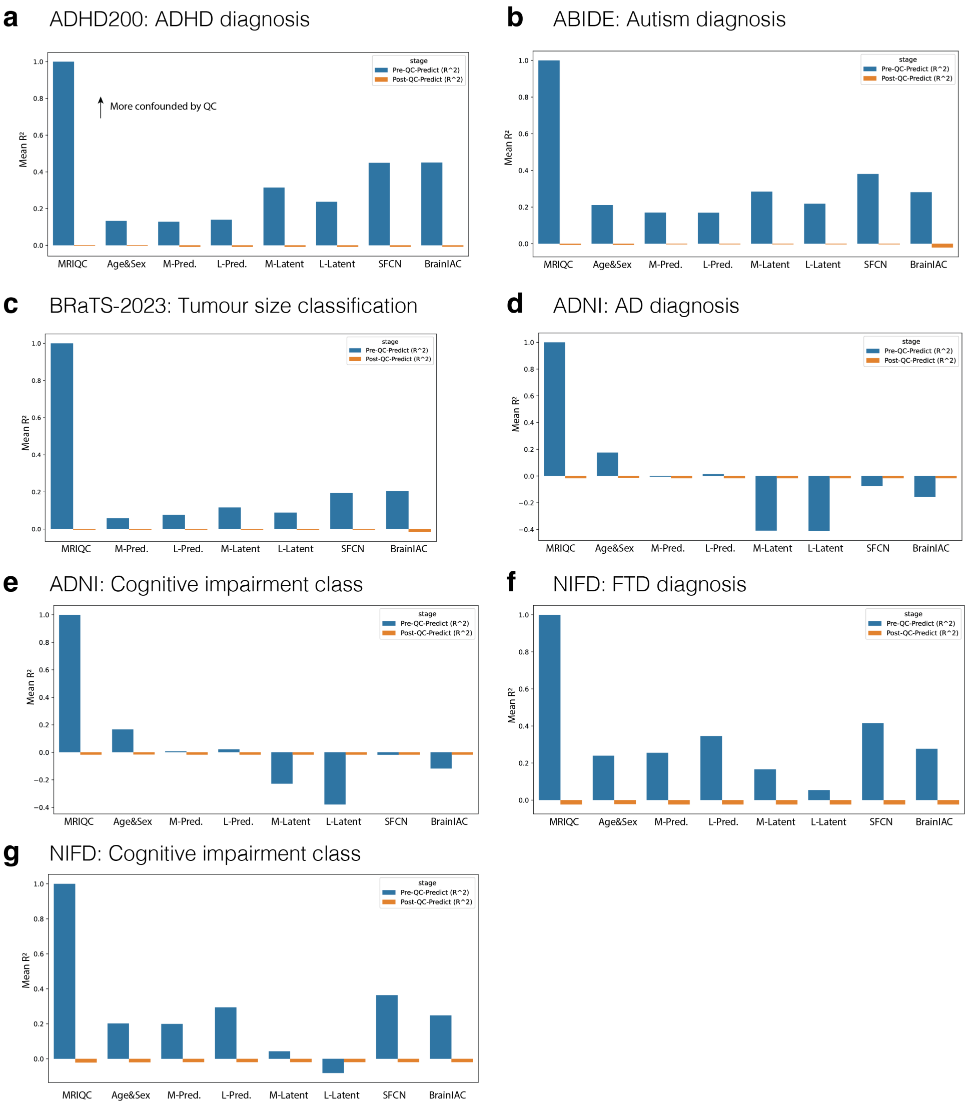


**Supplementary Fig. S8 | Pre- and post- residualization prediction of QC artefacts.** Mean cross-fold $R^2$ values from predicting quality control (QC) features using model latent representations before (blue) and after (orange) residualization within the double machine learning pipeline. Pre-residualization $R^2$ reflects linear predictability of QC confounds from each model's latent features (x-axis); post-residualization $R^2$ reflects the same after confound variance removal. M-Pred, L-Pred, M-Latent, and L-Latent denote four NeuroFM variants (Pred: predictor-head features; Latent: encoder-extracted features; M/L: model scale). NeuroFM latent features showed consistently lower QC confounding than SFCN or BrainIAC across cohorts. **a,** ADHD200. **b,** ABIDE. **c,** BraTS-2023. **d,** ADNI (diagnostic classifications). **e,** ADNI (cognitive assessment scores at time of scanning). **f,** NIFD (diagnostic classifications). **g,** NIFD (cognitive assessment scores at time of scanning).

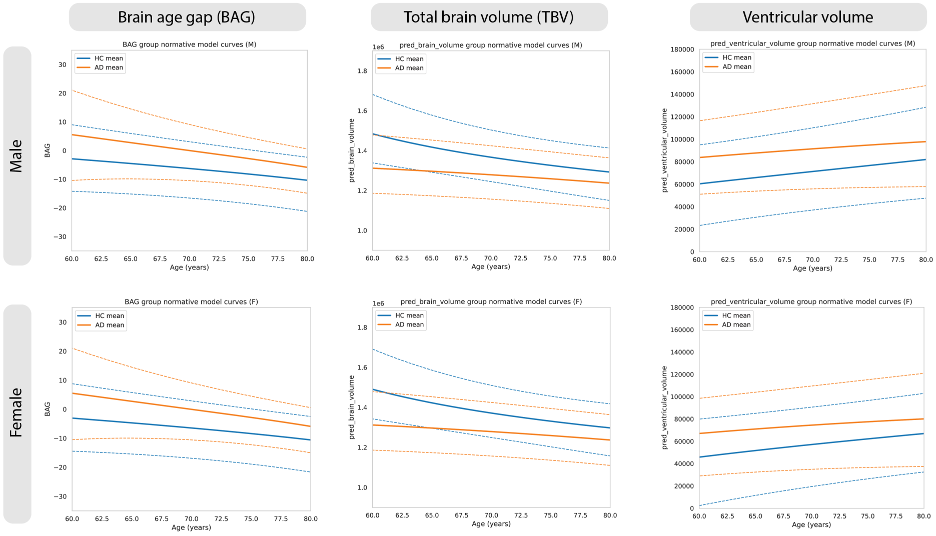


**Supplementary Fig. S9 | ADNI brain age gap and normative morphometric trajectories.** Normative models independently were fitted to three neuroimaging biomarkers derived from cross-sectional ADNI data: NeuroFM-predicted brain age gap (BAG), total brain volume (TBV), and ventricular volume. Results are stratified by sex (males, top row; females, bottom row) and Alzheimer's disease (AD) diagnostic status at time of scan (blue, cognitively normal; orange, AD-positive). TBV shows the expected age-associated decline and ventricular volume shows the expected age-associated expansion. BAG exhibits a decreasing mean trajectory with age, consistent with the known tendency of age-prediction models to regress toward the mean at the extremes of the training age distribution, producing apparent underestimation in older individuals. In both sexes and across all three biomarkers, the normative mean trajectory differs markedly between cognitively normal individuals and those with an AD diagnosis, indicating that each biomarker captures disease-related deviation from healthy ageing.

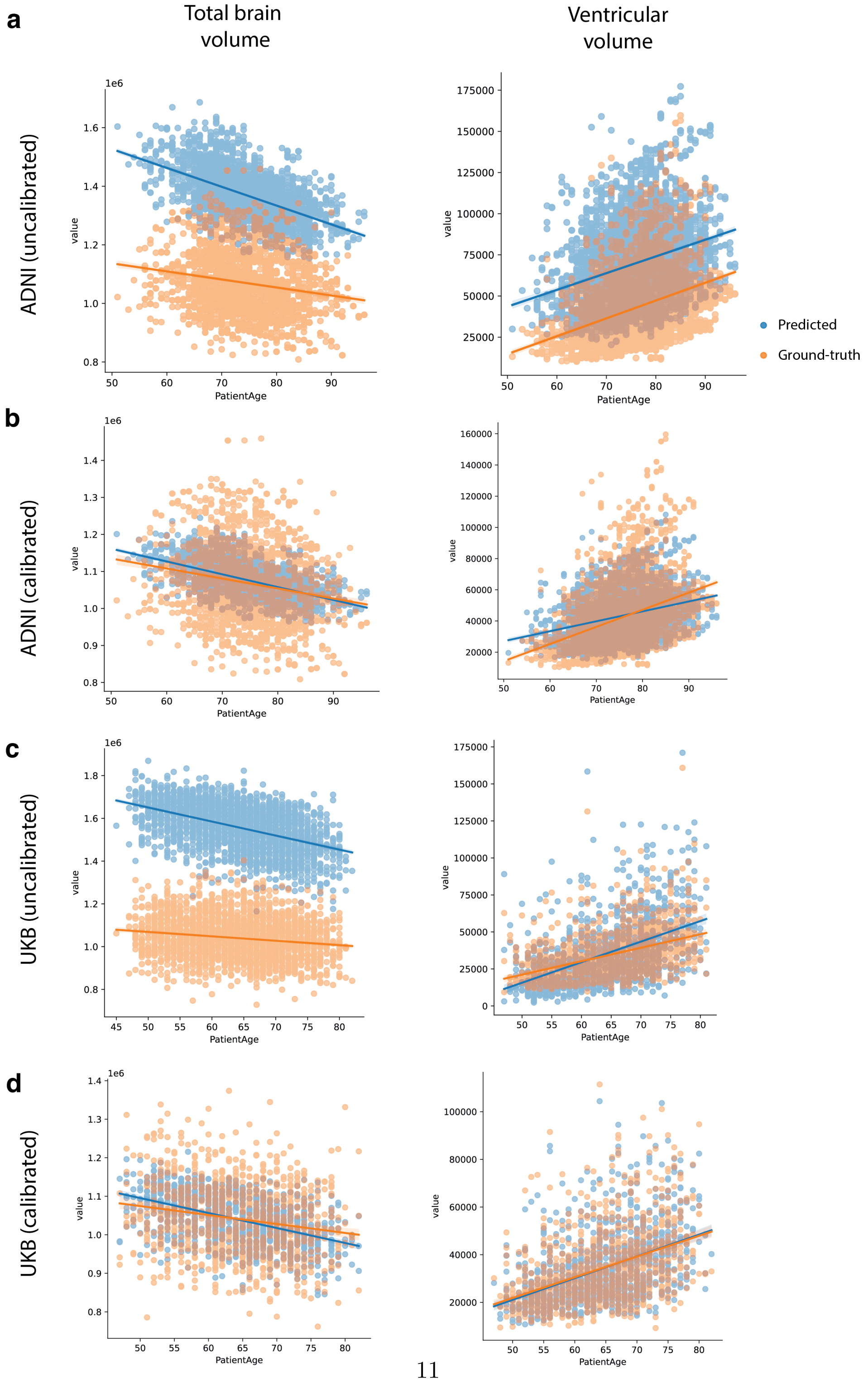




**Supplementary Fig. S10 | NeuroFM-M brain volume predictions compared to FreeSurfer ground-truth estimates across ADNI and UK Biobank cohorts.** Scatter plots show predicted (blue) and FreeSurfer-derived ground-truth (orange) estimates of total brain volume (left column) and lateral ventricular volume (right column) as a function of participant age, with linear regression trends overlaid for each. Results are shown before and after linear calibration; the calibration model was fit on a held-out 10% of volumes with subjects assigned exclusively to either the calibration or evaluation set to prevent data leakage. The substantial reduction in MAE following calibration is consistent with a systematic linear offset between predicted and ground-truth volumes, rather than errors in capturing age-related trajectories. In ADNI, calibration reduced MAE from 300,571 to 79,571 mm$^3$ for total brain volume and from 27,139 to 9,460 mm$^3$ for lateral ventricular volume. In the UK Biobank, MAE was reduced from 519,111 to 59,996 mm$^3$ for total brain volume and from 6,461 to 3,321 mm$^3$ for lateral ventricular volume. **a,** ADNI, uncalibrated. **b,** ADNI, calibrated. **c,** UK Biobank, uncalibrated. **d,** UK Biobank, calibrated.

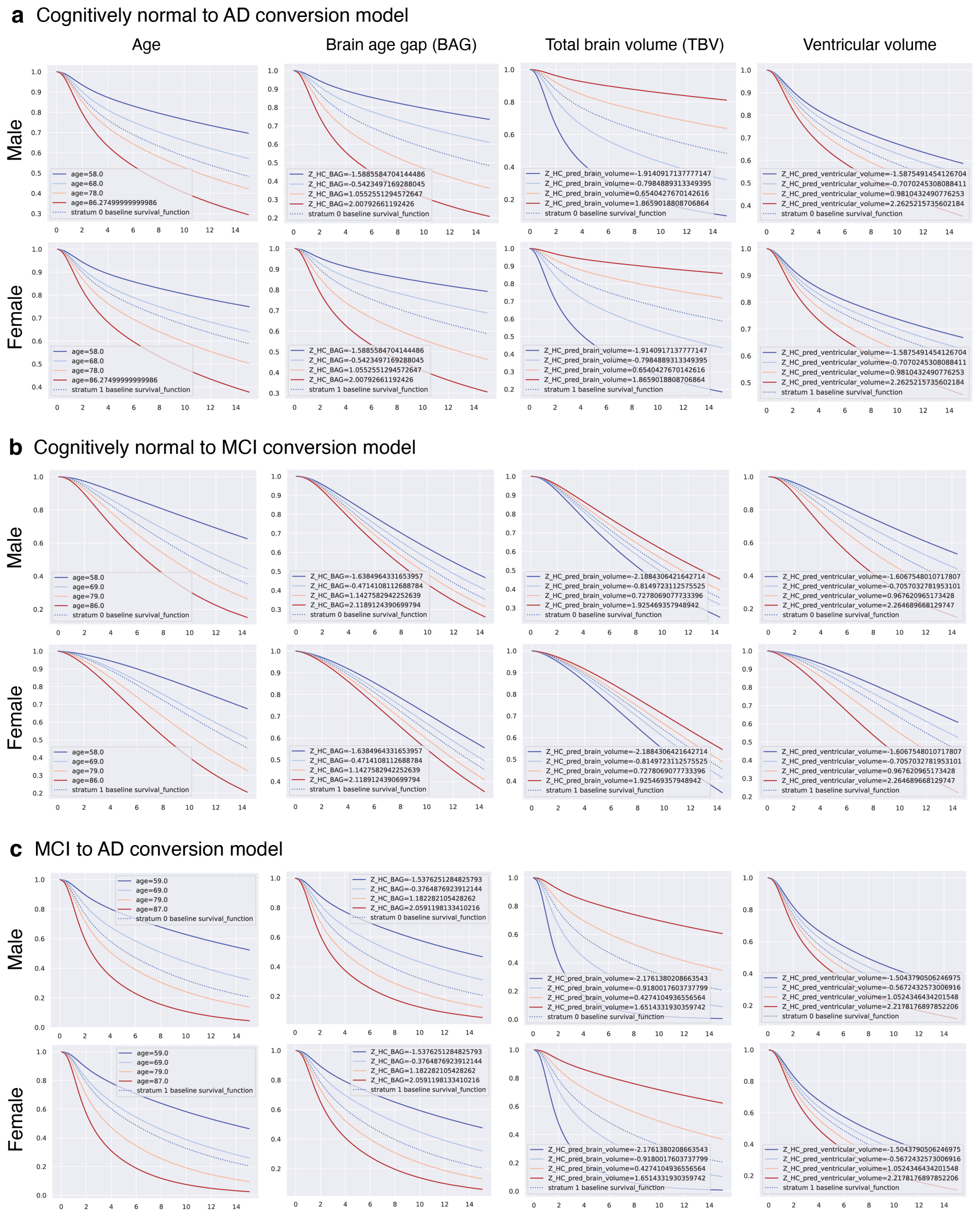


**Supplementary Fig. S11 | Cox proportional hazards partial effects plots stratified by covariate and sex.** Survival trajectories from Cox proportional hazards models plotted as a function of time since baseline visit. Each panel displays the partial effect of a single covariate, evaluated at the observed mean and at four additional values spanning the covariate distribution (coloured on a blue-to-red gradient from low to high). Columns correspond to the four covariates of interest: age, brain age gap (BAG), total brain volume (TBV), and ventricular volume. Because Cox models were fitted separately for each sex, male and female results are presented in separate rows. Three independent models were fitted to characterize progression risk under distinct clinical transitions. **a,** Cognitively normal (CN) to Alzheimer's disease (AD) conversion model. **b,** CN to mild cognitive impairment (MCI) conversion model. **c,** MCI to AD conversion model. Note that model **b** was excluded from the joint longitudinal-survival analysis reported in the main text, where only models **a** and **c** were employed.

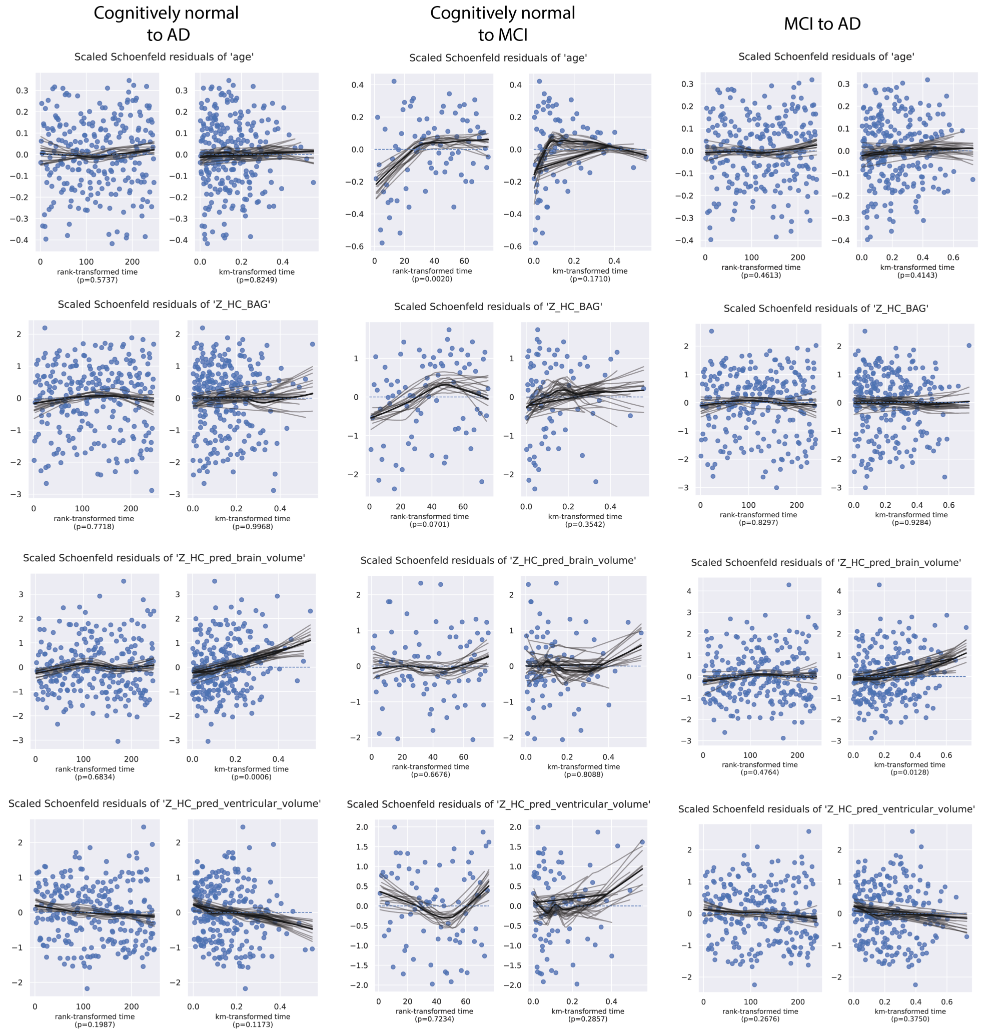


**Supplementary Fig. S12 | Assessment of proportional hazards assumptions for Cox regression models.** Scaled Schoenfeld residuals plotted against rank-transformed (left) and Kaplan-Meier-transformed (right) time axes for each covariate in three independently fitted Cox proportional hazards models characterizing distinct clinical transitions: cognitive normal (CN) to Alzheimer's disease (AD) conversion (left), CN to mild cognitive impairment (MCI) conversion (middle), and MCI to AD conversion (right). The CN-to-MCI model (middle) was excluded from the joint longitudinal-survival analysis reported in the main text. Grey marks indicate individual scaled Schoenfeld residuals at each event time; solid black lines represent loess-smoothed fits. Dashed lines indicate zero. P-values for non-proportionality (Grambsch-Therneau test) are shown for each covariate. Covariates are organized by row of plots. Although brain volume yields a statistically significant non-proportionality test in the models used for the main analysis, the residuals trend is flat under rank transformation and the apparent increase under KM-transformed time is confined to a sparsely sampled right tail, consistent with a right-censoring artifact rather than a genuine time-varying effect.

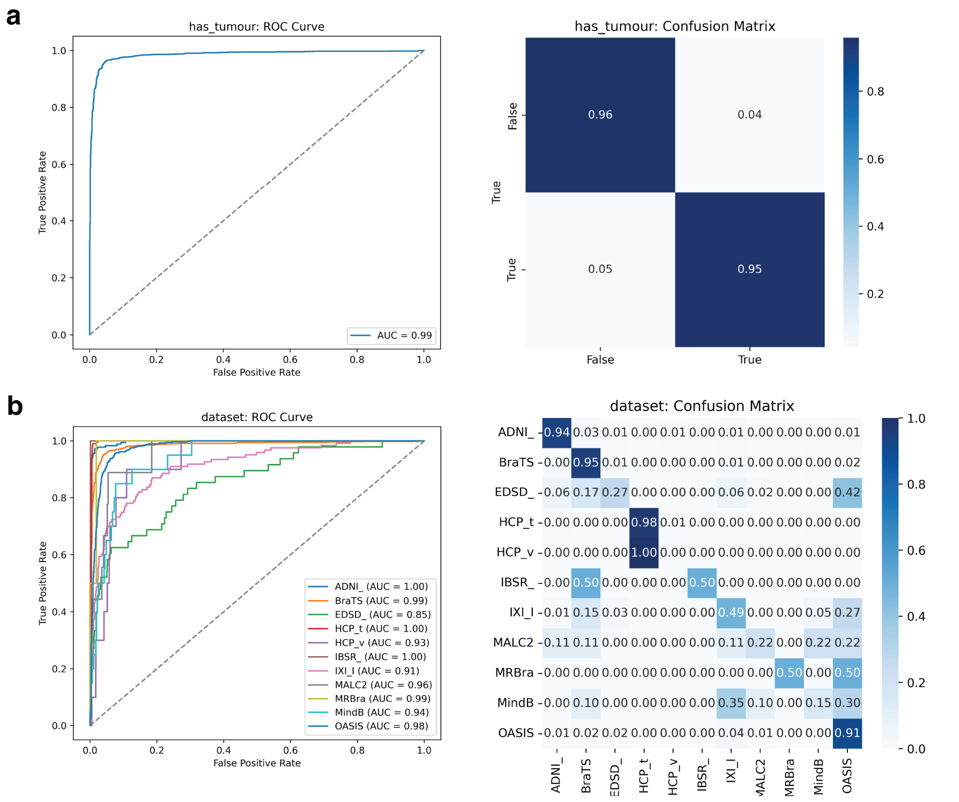


**Supplementary Fig. S13 | Multi-cohort confounding in tumour presence classification.** Tumour presence classification using external non-tumour cohorts as controls was initially attempted but abandoned due to model shortcutting on cohort-specific image artefacts rather than tumour-related signal. The main text therefore employed a within-BraTS-2023 binary classification of tumour volume (high vs. low) as a proxy for tumour burden, circumventing the absence of non-tumour control samples in that dataset. **a,** ROC curve for binary tumour presence classification (tumour vs. non-tumour controls from external cohorts), yielding an AUC of 0.99. Confusion matrix shown (right) demonstrates near-perfect separation, which we attribute to cohort-specific confounds rather than genuine tumour signal. **b,** To characterize the confounding effect, linear probing was applied to classify each sample by its source cohort across all 11 datasets. ROC curves are shown per cohort, with a minimum AUC of 0.85 and all remaining cohorts exceeding AUC = 0.90, confirming that cohort identity is strongly recoverable from the learned representations. Confusion matrix for cohort-level prediction shown (right). Together, these results indicate that the high tumour classification performance in **a** is driven by cohort-level imaging artefacts, motivating the use of a single-cohort tumour volume threshold design in the main analysis.

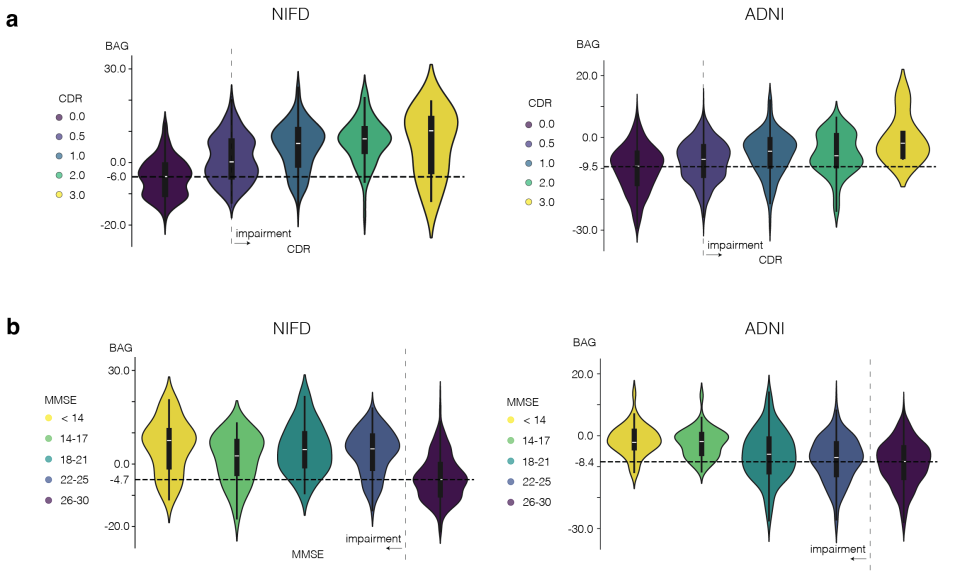


**Supplementary Fig. S14 | Uncorrected BAG relationships with cognition.** To make our BAG cognition analyses consistent with the classification tasks, predicted BAG values (shown in-text in Fig. 3) were residualized using QC values and age and sex. The uncorrected results are shown here. **a,** Uncorrected BAG in NIFD and ADNI shows significant mean predicted BAG differences compared the healthy group, increasing with impairment measured by the CDR score. **b,** Using MMSE, similar predicted BAG differences were observed in both datasets.

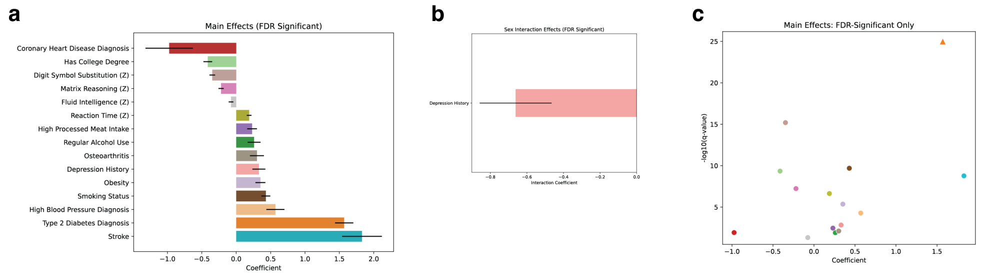


**Supplementary Fig. S15 | Clinical, lifestyle and cognitive associations with brain age gap and their moderation by sex using a pre-trained model without UK Biobank fine-tuning.** Whereas main-text Fig. 4 evaluates NeuroFM brain age associations after fine-tuning on the UK Biobank for separate T1-weighted and T2-weighted sequence analysis, here NeuroFM was applied as a frozen predictor of participant age without domain-specific adaptation. Inference was performed on the full T1-weighted UK Biobank sample (n = 43,943; 48,452 T1w volumes). All coefficients were corrected for multiple comparisons using the Benjamini–Hochberg false discovery rate (FDR) method. **a,** Standardized regression coefficients ($\beta\pm$ s.e.m.) for main effects on brain age gap (BAG; predicted minus chronological age). Higher BAG was most strongly associated with history of stroke, type 2 diabetes, high blood pressure and smoking status. **b,** Sex-interaction coefficients ($\beta\pm$ s.e.m.) for variables showing significant moderation by sex. Depression history showed a larger BAG association in females. Notably, coronary heart disease exhibited a strong negative interaction with BAG, a pattern absent in the fine-tuned model (main-text Fig. 4), which may reflect survivor bias or cardioprotective medication effects, though further investigation is needed. **c,** Volcano plot displaying effect size (x axis) against statistical significance (y axis) for main effects; capped values are denoted by triangles. Overall, the direction and ranking of main effects were broadly consistent with the fine-tuned model (main-text Fig. 4), though effects were generally smaller in magnitude and fewer reached statistical significance, consistent with the absence of domain-specific fine-tuning reducing the model's sensitivity to clinically relevant variation within this cohort.

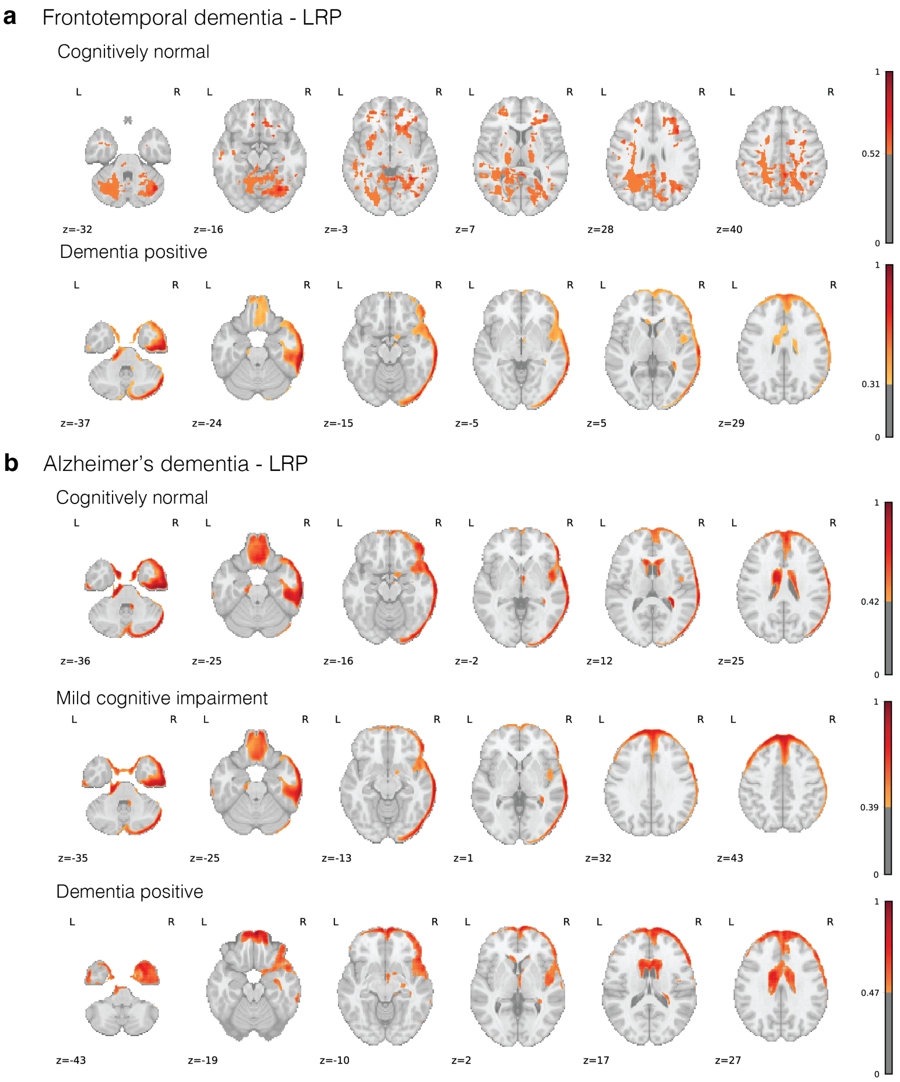


**Supplementary Fig. S16 | LRP results on frontotemporal and Alzheimer's dementia binary classification tasks. a,** Frontotemporal dementia LRP highlight results from NeuroFM. Cognitively normal predictions demonstrate anatomical plausibility. Dementia-positive predictions show primary highlighting near the outer CSF boundary, likely demonstrating movement-related confounds. **b,** Alzheimer's dementia LRP highlight results. While some highlighted regions in certain cases are plausible (ventricles, etc.), intensity near outer CSF boundary likely demonstrates movement confounds which were used as a prediction shortcut during finetuning.

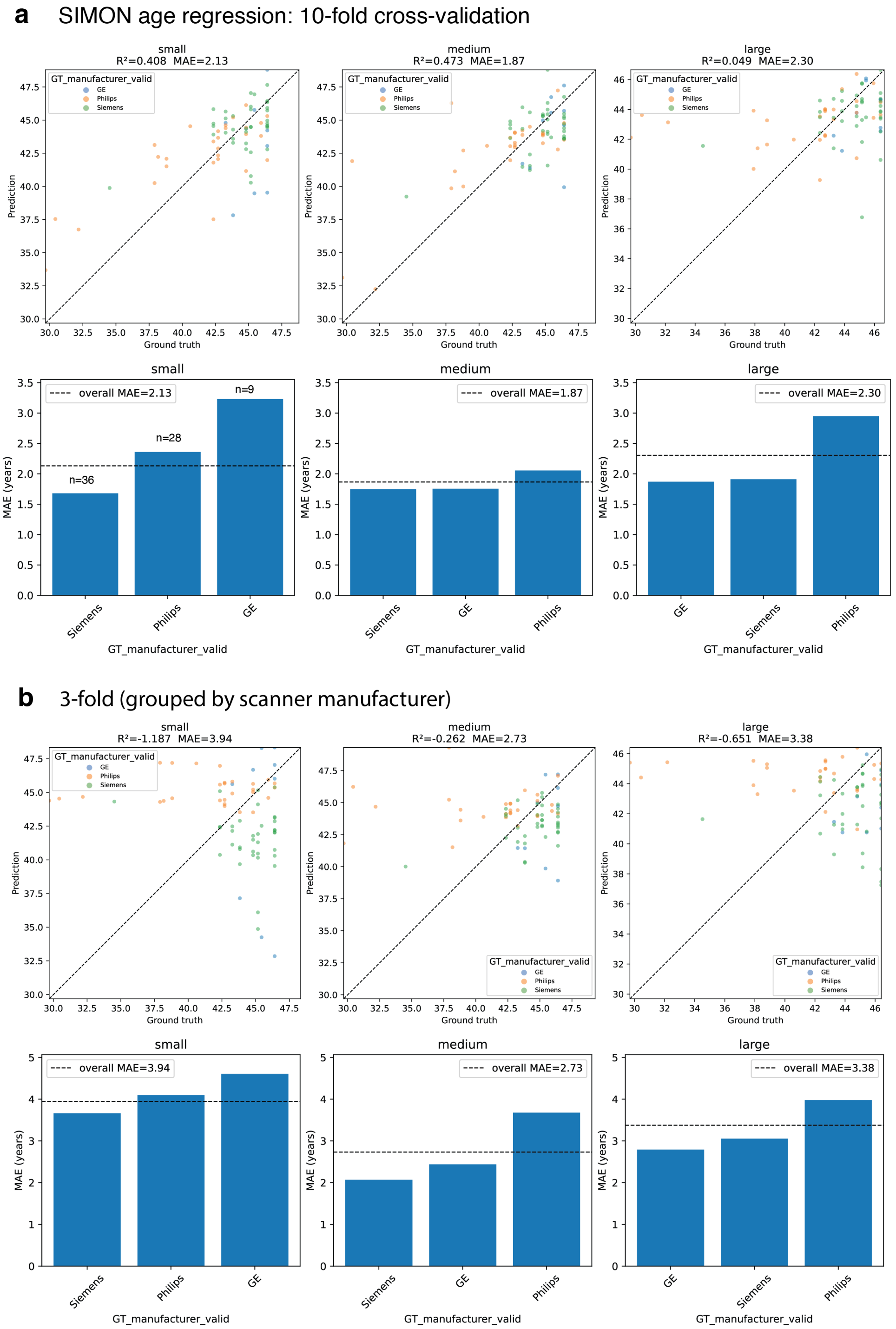




**Supplementary Fig. S17 | Brain age analysis on the SIMON dataset.** Linear regression models trained on NeuroFM representational features predicted chronological age, evaluating two generalization settings: cross-site transfer across a highly heterogeneous cohort ($n = 73$ scans; 33 acquisition sites; 15 scanner models) and out-of-distribution age prediction, as NeuroFM was pre-trained exclusively on individuals aged $\geq 45$ years while over half of SIMON images fell below this threshold. **a,** Ten-fold cross-validation. Scatter plots (top) show out-of-fold predicted versus chronological age across three NeuroFM capacity variants (columns), coloured by scanner manufacturer; bar plots (bottom) show mean absolute error (MAE) by manufacturer. **b,** Scanner-stratified three-fold cross-validation, withholding all scans from one manufacturer per fold to assess cross-manufacturer generalization. Note that the dataset is manufacturer-imbalanced: Siemens 50%, GE 12%, Philips the remainder. Layout follows **a**.

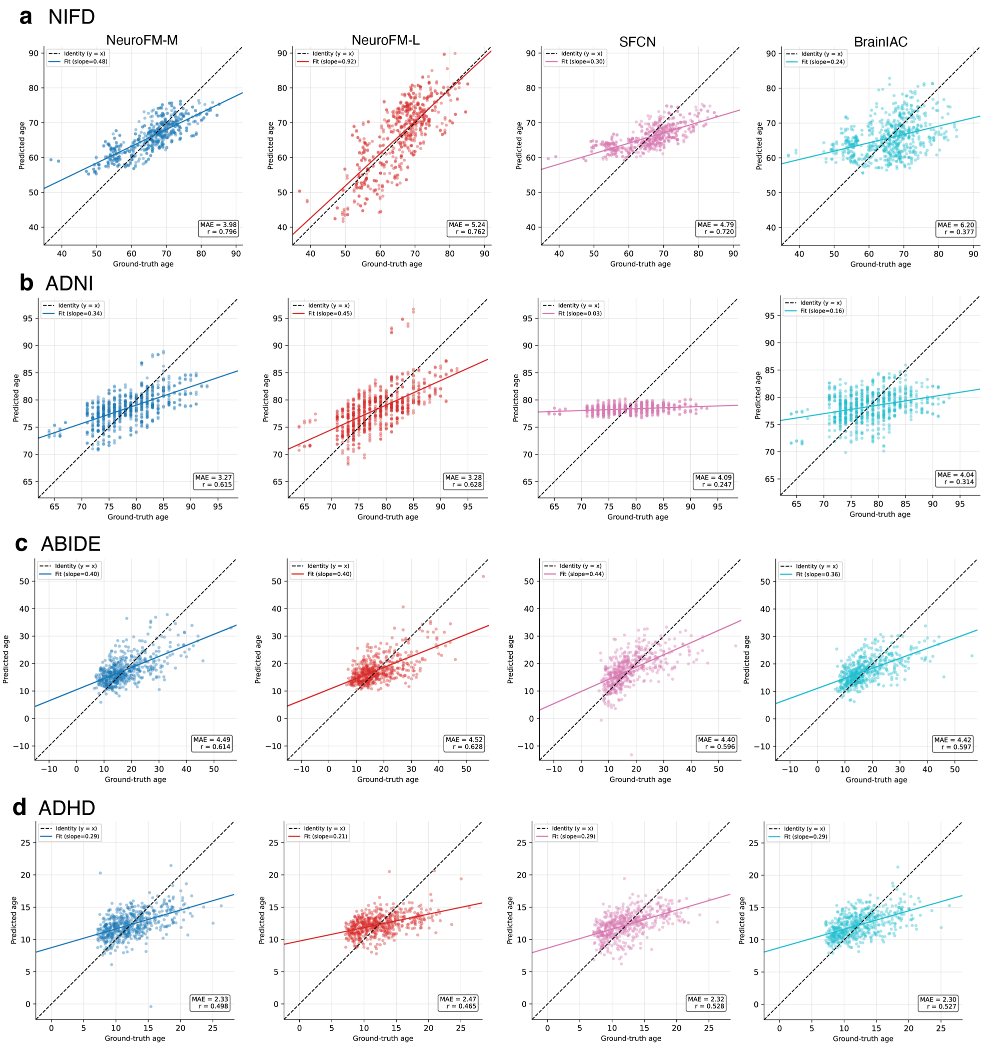


**Supplementary Fig. S18 | Brain age prediction bias across datasets and models.** Brain age was estimated for each individual using out-of-fold predictions from a linear regression model trained on the latent feature representations of each architecture, evaluated via cross-validation. Models are displayed left to right: NeuroFM-M, NeuroFM-L, SFCN, and BrainIAC. Scatter plots show predicted age versus chronological age for each cohort. **a,** NIFD cohort. **b,** ADNI cohort. **c,** ABIDE cohort, which includes both paediatric and adult participants. **d,** ADHD-200 cohort, which likewise includes both paediatric and adult participants.

### 2 Supplementary Tables

**Supplementary Table ST1 | MRIQC volume failure data.** Number of total volumes vs. the count of volumes that completed the MRIQC analysis pipeline. While overall success is near 100%, certain datasets like ADNI and LDM100k had a significant number of failures.

| Dataset | No. Volumes | No. Successful | No. Missing | % Missing |
|---|---|---|---|---|
| 1000_FCP | 7,820 | 7,494 | 326 | 4% |
| ABIDE | 1,010 | 1,010 | 0 | 0% |
| ADHD | 904 | 904 | 0 | 0% |
| ADNI | 17,320 | 3,098 | 14,222 | 82% |
| AOMIC | 1,911 | 1,911 | 0 | 0% |
| BRATS | 1,244 | 1,244 | 0 | 0% |
| EDSD | 290 | 290 | 0 | 0% |
| GLASGOW | 1,208 | 1,208 | 0 | 0% |
| HCP | 890 | 890 | 0 | 0% |
| IBSR | 18 | 18 | 0 | 0% |
| IXI | 569 | 569 | 0 | 0% |
| LDM100k | 99,994 | 1,860 | 98,134 | 98% |
| MRBrainS | 7 | 0 | 7 | 100% |
| MindBoggle101 | 92 | 92 | 0 | 0% |
| NIFD | 1,847 | 1,847 | 0 | 0% |
| OASIS3 | 2,633 | 2,633 | 0 | 0% |
| UK Biobank | 49,299 | 37,215 | 12,084 | 25% |

**Supplementary Table ST2 | Definitions and categories of MRIQC-derived quality metrics used in this study.** A mapping table for the MRIQC metric variables with their full names and category. '*' indicates where the metric variable has multiple subtypes, generally for white matter ("wm"), grey matter ("gm"), or CSF ("csf").

| Metric | Full Name | Category |
|---|---|---|
| cjv | Coefficient of joint variation | Tissue contrast / noise |
| cnr | Contrast-to-noise ratio | Tissue contrast |
| snr_* | Signal-to-noise ratio | Signal quality |
| snrd_* | SNR Dietrich | Signal quality |
| efc | Entropy focus criterion | Ghosting / motion |
| fber | Foreground-background energy ratio | Artifact / intensity |
| inu_* | Intensity non-uniformity | Bias field |
| fwhm_avg | Full-width half maximum | Smoothness |
| icvs_* | Intracranial volume fraction | Tissue composition |
| qi_2 | QI2 | Artifact detection |
| summary_*_mean | Tissue intensity mean | Intensity distribution |
| wm2max | White-matter-to-max ratio | Tissue contrast |

**Supplementary Table ST3 | Method 6 data after diagnostic and MRIQC (Method 12) removals.** Right column shows the final train set size (listed as the number of volumes) used in our Method 6 experiments. The total was calculated after removing the volumes that failed MRIQC analysis or did not have diagnostic data needed for classification. Note that there is some overlap (some volumes lack both diagnostic data and MRIQC results).

| Dataset | Total Vols | No. Lacking Diagnostic Data | No. Lacking MRIQC Results | Final No. Train Subjects | Final No.Train Volumes |
|---|---|---|---|---|---|
| ABIDE | 1,010 | 106 | 0 | 904 | 904 |
| ADHD | 904 | 24 | 0 | 880 | 880 |
| ADNI | 17,320 | 4,295 | 14,222 | 265 | 2,450 |
| BRATS | 1,244 | — | 0 | 1,127 | 1,244 |
| NIFD | 1,847 | 396 | 0 | 259 | 1,451 |

**Supplementary Table ST4 | Training and testing data for quality metric prediction (Method 12).** The quantity, per dataset, of volumes used for training and testing the quality metric prediction task. Splits were grouped by subject ID, so there was no repeat subject leakage between training and testing.

| Dataset | No. Test | No. Train |
|---|---|---|
| 1000_FCP | 0 | 7,494 |
| ABIDE | 810 | 200 |
| ADHD | 704 | 200 |
| ADNI | 1,118 | 1,982 |
| AOMIC | 0 | 1,911 |
| BRATS | 1,022 | 222 |
| EDSD | 0 | 290 |
| GLASGOW | 0 | 1,208 |
| HCP | 0 | 890 |
| IADNI | 0 | 98 |
| IBSR | 0 | 18 |
| IXI | 0 | 569 |
| LDM100k | 1,660 | 200 |
| MindBoggle101 | 0 | 92 |
| NIFD | 778 | 1,071 |
| OASIS3 | 0 | 2,633 |
| UKB | 34,993 | 3,008 |
| **TOTAL** | **41,085** | **22,086** |

**Supplementary Table ST5 | Augmentation specifications.** The parameters and probability for each augmentation used in our NeuroFM pretraining regime. We used blur, downscale, and ghosting from Perez-Garcia et al. (2021), and motion from Shaw et al. (2019). Some augmentations are applied independently, so probabilities do not sum to 1.

| Group | Augmentation | Prob | Parameters |
|---|---|---|---|
| **Geometrical** | Translation | 1/3 | 3 axes; shift ±20 voxels; zero padded |
| | Rotation | 1/3 | 3 axes; ±10 degrees; zero padded |
| **Intensity-based (Noise)** | Blur | 1/10 | Limit: 3 |
| | Salt & pepper | 1/40 | Amount: 0.01; Salt: 0.2 |
| | Gaussian | 1/30 | Amount: 0.2 |
| | Downscale | 1/8 | Scale: 0.25–0.75 |
| | Gamma | 1/20 | Clip: 0.025 |
| | Contrast | 1/10 | Alpha: 0.5–3.0 |
| **Intensity-based (Artefacts)** | Ghosting | 1/15 | Max repetitions: 4 |
| | Slice Spacing | 1/8 | Axial plane only; Spacing: 2–5 mm |
| | Inhomogeneity | 1/8 | See Svanera et al. (2021) for details |
| | Field Bias | 1/12 | Num cycles: 5; Scale factor: 2 |
| | Motion | 1/15 | Rotation: ±10 degrees; Translation: ±5 voxels; Transforms: 2 |

**Supplementary Table ST6 | NeuroFM pretraining multi-task performance.** Each model variant was trained independently in a supervised fashion to predict participant age, brain volume, ventricle volume, and sex. For the regression tasks, $R^2$ was used as a closeness-of-fit estimate to compare across models. Reported accuracy is balanced accuracy. Bold values indicate the best performance per column.

| Model Variant | # Parameters | Age ($R^2$) | Brain Volume ($R^2$) | Ventricle Volume ($R^2$) | Sex (Accuracy) |
|---|---|---|---|---|---|
| S | 484,829 | 0.83 | **0.96** | 0.97 | 0.99 |
| M | 6,535,877 | **0.86** | 0.95 | **0.97** | 0.99 |
| L | 10,815,109 | 0.85 | 0.95 | 0.96 | 0.99 |

**Supplementary Table ST7 | NeuroFM pretraining hyperparameters.** Hyperparameters for model variants S, M, and L. All models used ELU activation, batch normalization, Adam optimizer, exponential LR decay ($\gamma = -0.01$), and float16 precision. Augmentation probabilities are identical across variants. A dash (—) indicates the feature was disabled or not used.

| **Parameter** | **S** | **M** | **L** |
|---|---|---|---|
| ***Architecture*** | | | |
| Convolutional layers | 5 | 7 | 8 |
| Initial filters | 32 | 32 | 32 |
| Conv block | Bottleneck | Bottleneck | Bottleneck |
| Identity layers | — | 5 (from layer 2) | 5 (from layer 2) |
| Final stage | Avg. pooling | Avg. pooling | Dense |
| Neck layers (× size) | — | 2 × 256 | 2 × 512 |
| Dense predictor size | — | 128 | 256 |
| Dropout rate | 0.10 | 0.25 | 0.25 |
| ***Training*** | | | |
| Learning rate | $4 \times 10^{-4}$ | $7 \times 10^{-5}$ | $6 \times 10^{-5}$ |
| Batch size | 14 | 10 | 8 |
| Age loss weight | 0.30 | 0.30 | 0.35 |
